# Design and evaluation of mobile monitoring campaigns for air pollution exposure assessment in epidemiologic cohorts

**DOI:** 10.1101/2021.04.21.21255641

**Authors:** Magali N. Blanco, Annie Doubleday, Elena Austin, Julian D. Marshall, Edmund Seto, Timothy Larson, Lianne Sheppard

## Abstract

Mobile monitoring campaigns to estimate long-term air pollution levels are becoming increasingly common. Still, many campaigns have not conducted temporally-balanced sampling, and few have looked at the implications of such study designs for epidemiologic exposure assessment. We carried out a simulation study of fixed-site air quality monitors to better understand how different mobile monitoring designs involving short-term stationary measurements at fixed locations impact the resulting exposure surfaces. We used Monte Carlo resampling to simulate three archetypal monitoring designs using oxides of nitrogen (NOx) monitoring data from 69 regulatory sites in California: a year-around Balanced Design that sampled during all seasons of the year, days of the week, and all or various hours of the day; a temporally reduced Rush Hours Design; and a temporally reduced Business Hours Design. We evaluated the performance of each design’s land use regression prediction model. The Balanced Design consistently yielded the most accurate annual averages; while the reduced Rush Hours and Business Hours Designs generally produced more biased results. A temporally-balanced sampling design is crucial for mobile monitoring campaigns aiming to assess accurate long-term exposure in epidemiologic cohorts.

**Synopsis:** Air pollution mobile monitoring campaigns rarely conduct temporally balanced sampling. We show that this results in biased annual average exposure estimates.

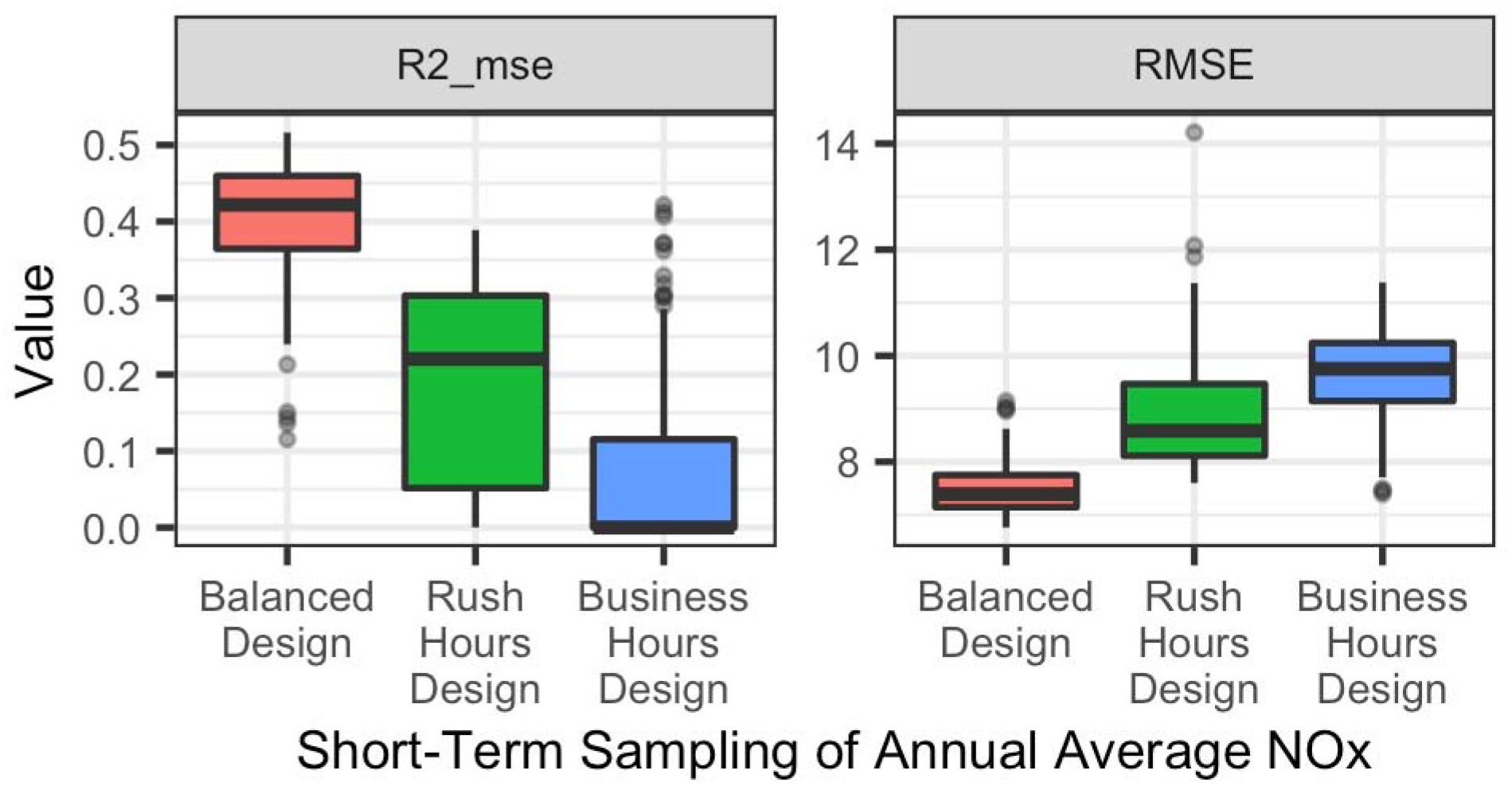

## 1 Introduction

A large body of evidence links long-term exposure to air pollution to adverse health effects in humans, including mortality from cardiovascular outcomes and lung cancer.^1–6^ An increasing number of studies are using mobile monitoring campaigns to assess average long-term air pollutant levels.^7–13^ Mobile monitoring campaigns typically equip a vehicle with air monitors and collect short-term samples while in motion (non-stationary sampling) and/or while stopped (stationary sampling). The focus of this analysis is on the latter mobile monitoring design. A single monitoring platform can be used to collect samples at many specified locations within a relatively short period of time, making it a time and cost-efficient sampling approach. Mobile campaigns are particularly well-suited for multi-pollutant monitoring of less frequently monitored traffic-related air pollutants that require expensive instruments or instruments that need frequent attention during the sampling period.

And while a few studies have investigated the number of sampling locations and repeat samples needed to improve the resulting exposure surfaces from mobile monitoring campaigns,^14, 15^ to the best of our knowledge, none have considered the importance of conducting temporally-balanced sampling when the goal is estimation of a long-term average for epidemiologic application. This is particularly relevant since many pollutants, particularly those related to traffic, experience strong diurnal and seasonal concentration trends.^16, 17^ Collecting limited or unbalanced sampling may thus be sufficient to answer questions surrounding peak concentrations or source identification, but it may produce biased long-term estimates and be inadequate for epidemiologic applications.^22^ In general, many mobile monitoring campaigns have been short, lasting from a few weeks to months and with few repeat visits to each location spanning one to three seasons.^9, 18–20^ Most campaigns have conducted sampling during weekday business or rush hours, ignoring the surrounding hours, when air pollution concentrations can be drastically different.^8, 17, 21^ Furthermore, short-term mobile monitoring campaigns often collect non-stationary (mobile) measurements, which can be much shorter in duration than stationary campaigns (e.g., a few seconds per road segment vs minutes or hours per stop location). It is an open question whether these shorter sampling times along with the platform’s increased proximity to immediate vehicle sources (e.g., in a traffic queue while stopped at a traffic signal) may produce more biased and less precise exposure surfaces when compared to short-term stationary monitoring.^12, 23–26^

The goal of this paper is to shed light on the temporal design of a short-term stationary mobile monitoring campaign for application to epidemiologic cohort studies. We carry out a set of simulation studies to better understand the role of mobile monitoring design on the prediction of annual average surfaces. We use existing monitoring data from California to compare the primary, annual site averages when all of the data are included to subsequent analyses utilizing subsets of the data. These data provide a unique opportunity to explore how short-term stationary sampling strategies can influence the resulting estimated annual-average concentration. Our analysis requires having a long-term, comprehensive set of measurement data, which therefore necessitates using fixed-site measurements rather than mobile measurements, to shed light on an aspect of study design for short-term stationary mobile monitoring.

## 2 Methods

### 2.1 Data

We simulate three sampling designs (see below) using hourly observations for oxides of nitrogen (NOx) collected during 2016 from regulatory Air Quality System (AQS) sites in California. NOx was selected since it is a spatially and temporally variable traffic pollutant with a strong diurnal pattern,^8, 27, 28^ and it is measured at many regulatory monitoring sites in California, providing a large enough dataset for this analysis.^29^

We included 69 of 105 California AQS sites that met various criteria (**Error! Reference source not found.**, SI Figure S3). First, sites needed to have readings at least 66% of the time (5,797/8,784 hourly samples; 2016 was a leap year). Second, sites needed to have sampling throughout the year, such that data collection gaps were a maximum of 45 days long. These two criteria are in line with other air quality work.^30^ Third, sites were required to have sampled for at least 40% of the time during various two-week periods that were used in two of our “common” designs (described below). This sample size ensured that we could sample during these periods without replacement. Fourth, sites were required to have positive readings (> 0 ppb) at least 60% of the time, thus ensuring that sites had sufficient variability in their concentrations and allowing us to model annual averages on the natural log scale. Finally, sites in rural and industrial settings (as determined by the US EPA)^31^ were excluded since these do not represent where the majority of people reside. The resulting sites were in both urban and suburban settings, in residential and commercial areas.

### 2.2 Sampling Designs

We conducted simulation studies to characterize the properties of three sampling designs (

Table 1, Supplementary Information [SI] Figure S1). Each design has a long- and a short-term sampling approach. Long-term approaches use all of the data that meet each design’s definition to estimate site annual averages and are analogous to traditional, fixed-site sampling approaches where sampling at a given location occurs over an extended period of time. Short-term approaches only collect 28 samples per site and are analogous to mobile monitoring campaigns that collect a few repeat samples per site. (The cut-off of 28 samples reflects our preliminary analyses showing that 28 hourly NOx samples are sufficient to estimate a site’s annual average within about 25% error or less [SI Figure S2].) Each design has multiple versions where samples are collected at slightly different times. The various design versions are intended to reflect the bias produced if only certain times are included in the measurements. We simulated each short-term sampling approach 30 times (Monte Carlo resampling), and hereafter refer to each of these simulations as a “campaign” since each represents a potential mobile monitoring study.

**Table 1.**
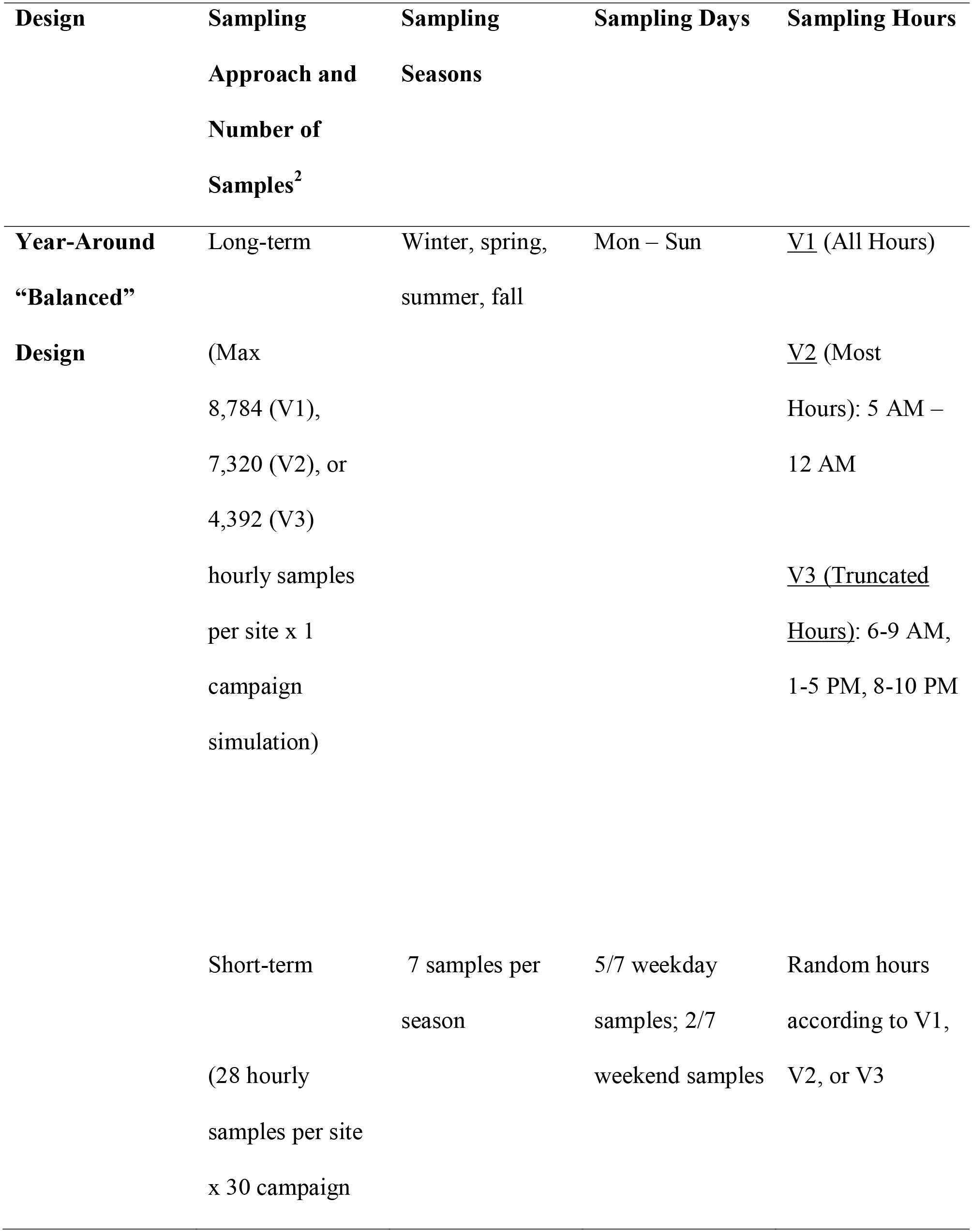
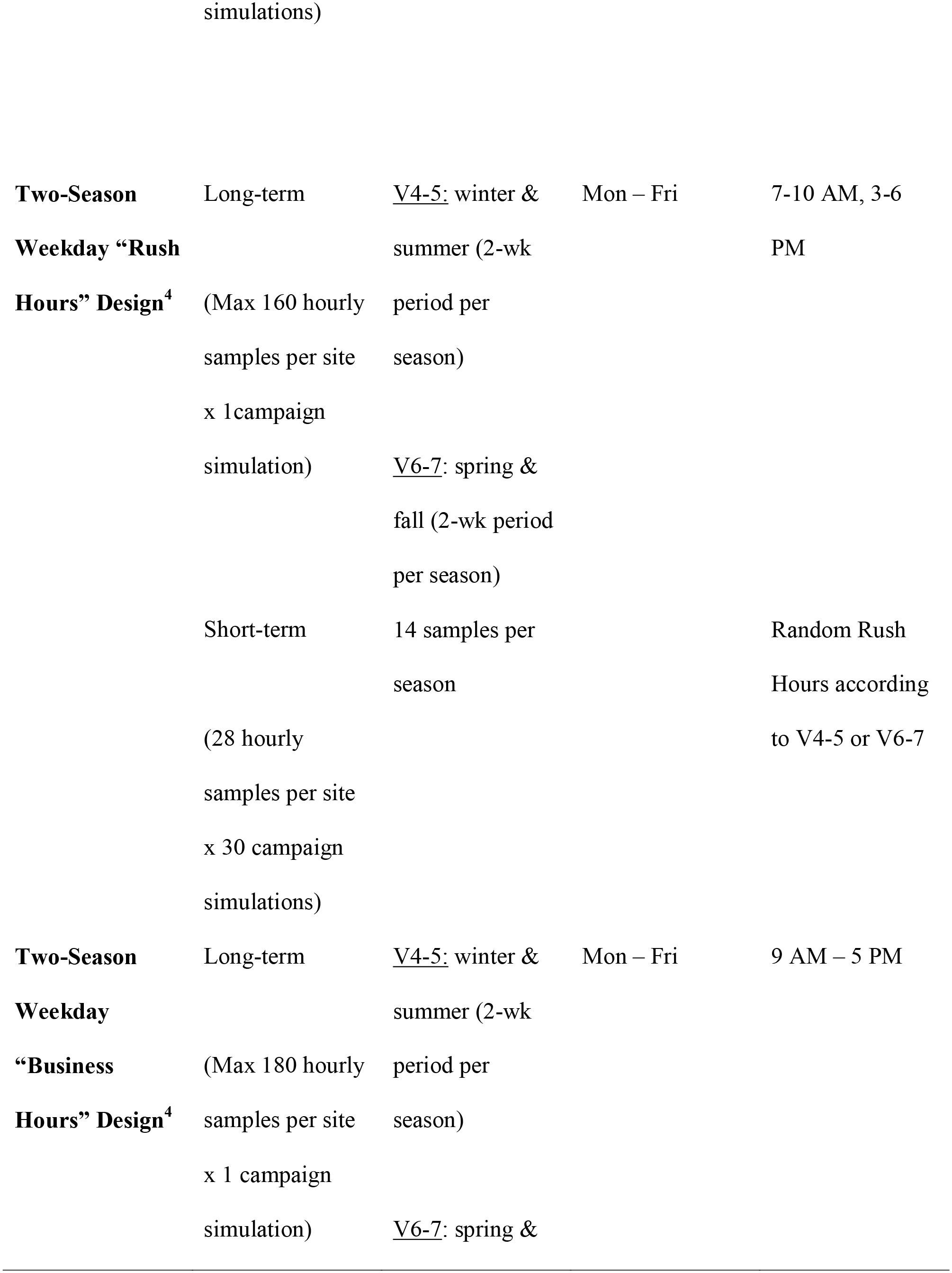
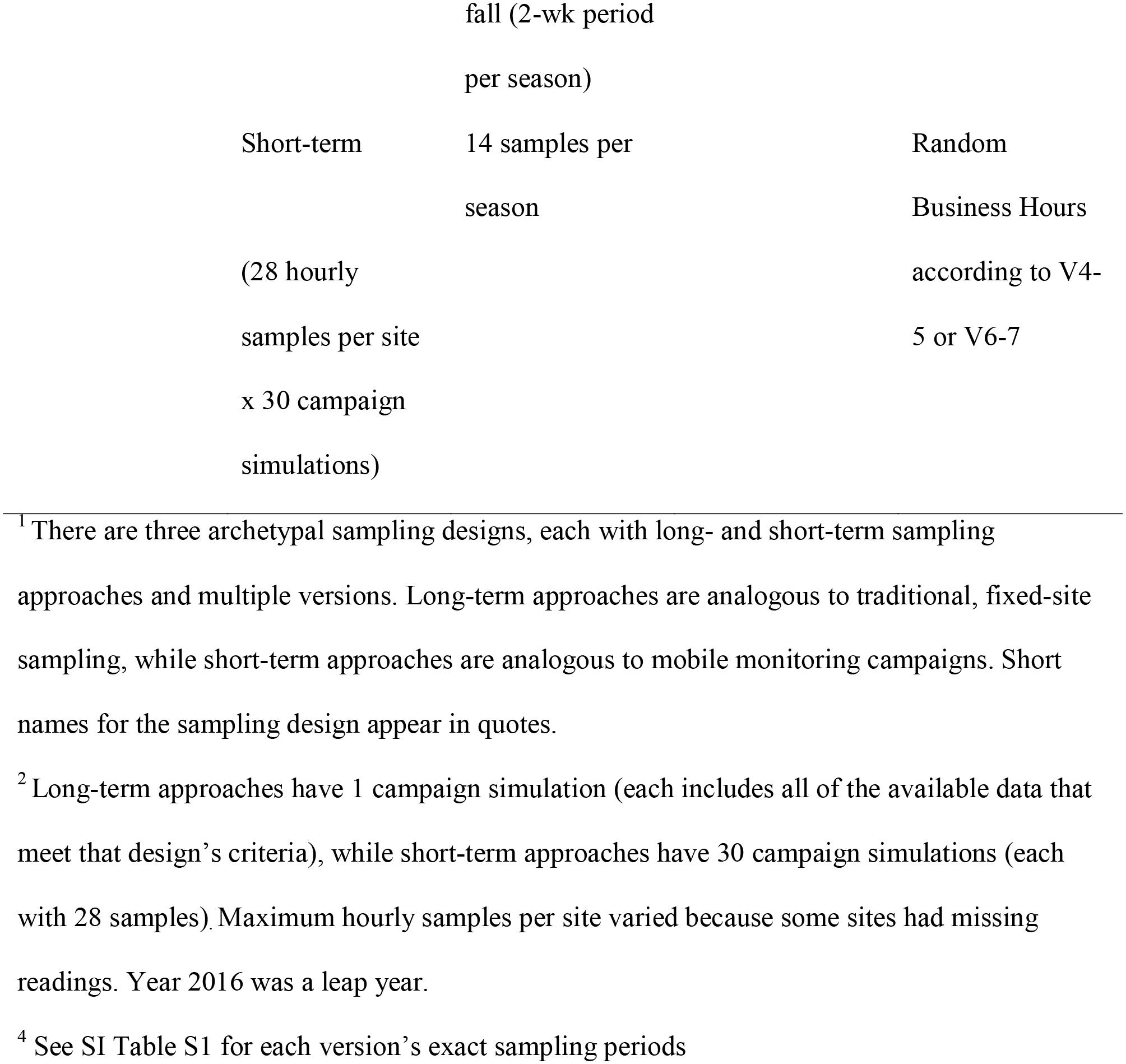
Simulated sampling designs used to estimate site annual averages^1^.

The Year-Around “Balanced” Design represents an “ideal” sampling scheme: sampling is conducted during all seasons, days of the week, and all or most hours of the day. Version 1 collects samples during all hours of the day. Versions 2-3 reduce the sampling hours to reflect the logistical constraints of executing an extensive campaign: samples occur during most hours of the day (5 AM – 12 AM only; “Version 2”) or during 6-9 AM, 1-5 PM and 8-10 PM (“Version 3”). Estimates from the long-term Balanced Design Version 1 are analogous to what might be collected from a traditional, year-around, fixed-site sampling scheme. For simplicity, we interchangeably refer to these as the “true” estimates or the “gold standard” hereafter, though we acknowledge that some error exists (e.g., due to missing hours or instrument accuracy).

The Two-Season Weekday “Rush Hours” and “Business Hours” Designs reflect common designs in the literature. Samples are collected either during summer and winter (Versions 4-5) or spring and fall (Versions 6-7). Sampling for each version occurs on weekdays during the same two-week period for all sites during each relevant season (See SI Table S1 for each version’s exact sampling periods). Sampling is restricted to the hours of 7-10 AM and 3-6 PM (Rush Hours Design) or 9 AM – 5 PM (Business Hours Design). The short-term approach collects 14 random samples during each season.

### 2.3 Prediction Models

We estimated unweighted site annual averages based on the data collected during each campaign. We log-transformed these before using them as the outcome variable in partial least squares (PLS) regression models, which summarized hundreds of geographic covariate predictors (e.g., land use, road proximity, and population density; see SI Table S2 for the covariates considered) into two PLS components (using the `plsr` function in the `pls` package in R). We evaluated the performance of each campaign using ten-fold cross-validated (CV) predictions on the native scale, incorporating re-estimation of the PLS components in each fold. The cross-validation groups were randomly selected and, importantly, fixed across all campaigns to allow for consistent model performance comparisons across design versions.

To best understand the role of design, we present results for annual average estimates, predictions, and model performance statistics. In descriptive analyses, we compare design-specific annual average estimates and predictions to the gold standard (long-term Balanced Design Version 1). We compare predicted site concentrations against predictions from the gold standard since epidemiologic air pollution studies often rely on predicted exposure, and the gold standard prediction represents the best possible prediction of annual-average concentrations that a study could hope to achieve. We complement this approach with model assessment evaluations of design-specific site predictions against two different references: an assessment against the true averages, and a traditional model assessment evaluation against the respective design-specific annual average estimates. The traditional assessment compares the predicted exposures to the observed site measurements from which they were derived. This allows us to document the quantities that would normally be available from modeling the data measured from any specific campaign. We summarize the model performance in terms of cross-validated mean squared error (MSE)-based R^2^ (R^2^_MSE_), regression-based R^2^ (R^2^_reg_), and root mean squared error (RMSE). R^2^_MSE_ assesses whether two sets of measurements such as estimates and predictions are the same (along the 1-1 line), and thus reflects both bias and variation around the one-to-one line (see SI Equations 1-3 for definitions). R^2^ _reg_, on the other hand, assesses whether observations are linearly associated (based on the best fit line though not necessarily the 1-1 line) and thus adjusts for bias and slopes different than one. R^2^_reg_ is defined as the squared correlation between two sets of measurements.

In sensitivity analyses, we repeated these simulations for nitrogen dioxide (NO_2_) and nitrogen monoxide (NO), adding a two ppb constant to all of the hourly NO readings before log-transforming to eliminate negative and zero concentration readings. Furthermore, we conducted NOx simulations for a subset of sites (N=17) within the Los Angeles (LA) and San Diego Counties, refitting PLS models to these sites alone. This region was meant to represent a potential area of interest for epidemiologic exposure assessment and one that could be more feasibly covered by a mobile monitoring campaign, though it had a reduced sample size.

*All analyses were conducted in R (v 3.6.2, using RStudio v 1.2.5033).^32^ SI Note S1 lists the R packages used. All map tiles were created by Stamen Design^33^ under CC BY 3.0,^34^ using data by OpenStreetMap^35^ under ODbL.*

## 3 Results

### 3.1 Hourly Readings

Sites (N=69) had on average (SD) of 8,090 (361) hourly readings, the equivalent of 337 (15) days of full sampling (See SI Table S3). Average (SD) hourly NOx concentrations were 16 (21) ppb (See SI Table S4). Sites had seasonal, daily, and hourly concentration patterns, with trends being more pronounced at some sites than others (See SI Figure S4-S6).

### 3.2 Annual Average Estimates

Across the 69 monitor locations, measured annual average concentrations (long-term Balanced Design Version 1), had a median (IQR) of 14 (10 - 21) ppb and ranged from 3-56 ppb. The short-term and long-term sampling approaches resulted in similar distributions of annual averages for different design versions. Figure 2 shows the long-term and a single short-term approach for each design. Overall, the long-term and short-term approach for each design version had very similar distributions. All of the Balanced Design versions resulted in only slight differences in their medians and IQRs. The Rush Hours Design versions generally resulted in slightly higher annual averages than the true averages, with some versions being more variable and having somewhat different distributions. The Business Hours Design versions resulted in annual averages that were generally lower than the true averages and less variable than the Rush Hours Design versions. See SI Table S5 for summary statistics. SI Figure S7 shows annual average estimates for all campaigns and pollutants.

**Figure 1.**
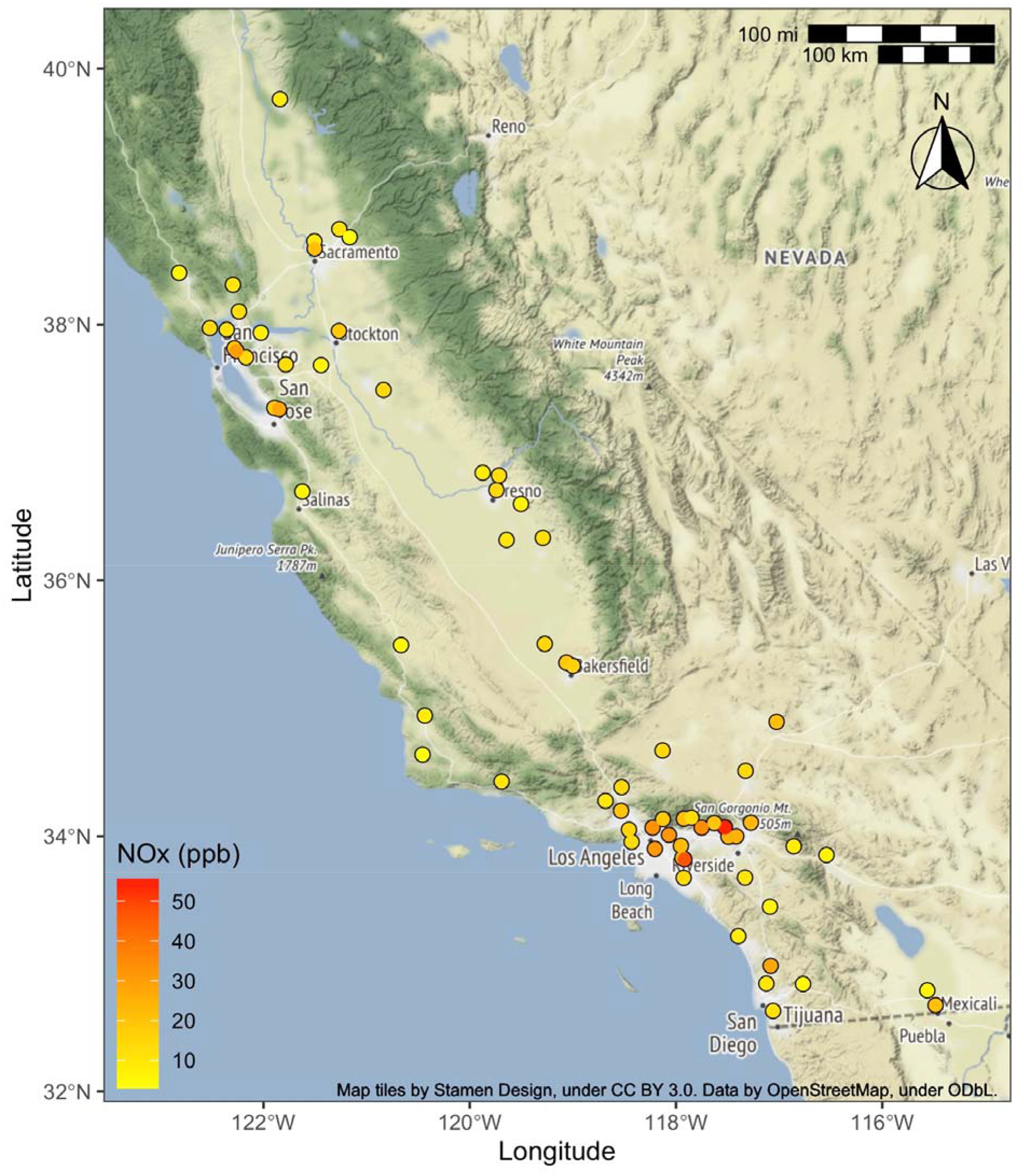
AQS sites included in this analysis (N=69) and their true annual average NOx measurements, as measured by the long-term Year-Around Balanced Design Version 1 (see Methods for details).

**Figure 2.**
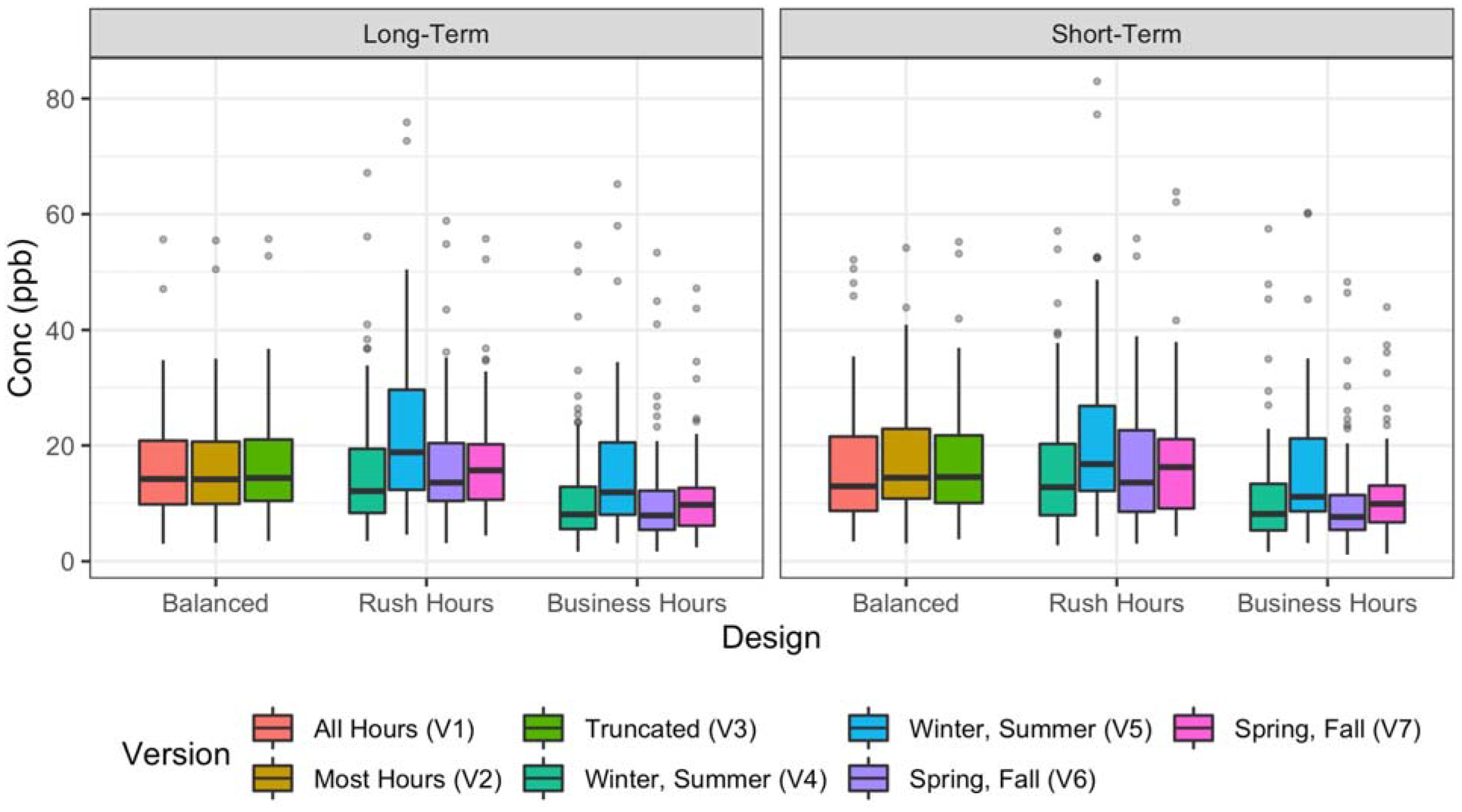
Distribution of NOx annual averages (N=69 sites) from different design versions. Showing the one campaign for each long-term approach and one example campaign for each short-term approach.

Figure 3 shows the site-specific distributions of annual averages across designs for short-term approaches relative to the true averages for a stratified random sample of 12 sites. Sites are stratified by whether their true mean concentration was in the low (<25^th^ percentile), middle (25^th^-75^th^ percentile) or high (>75^th^ percentile) concentration category. The variation of averages across campaigns increases with concentration in all designs. Site-specific averages are similar to the true averages for all Balanced Design versions while there were multiple sites from the Business Hours Design versions with averages systematically lower. The Rush Hours Design versions also had many biased averages, although the direction of the bias varied by site and design version. SI Figure S8 shows these biases for all sites.

**Figure 3.**
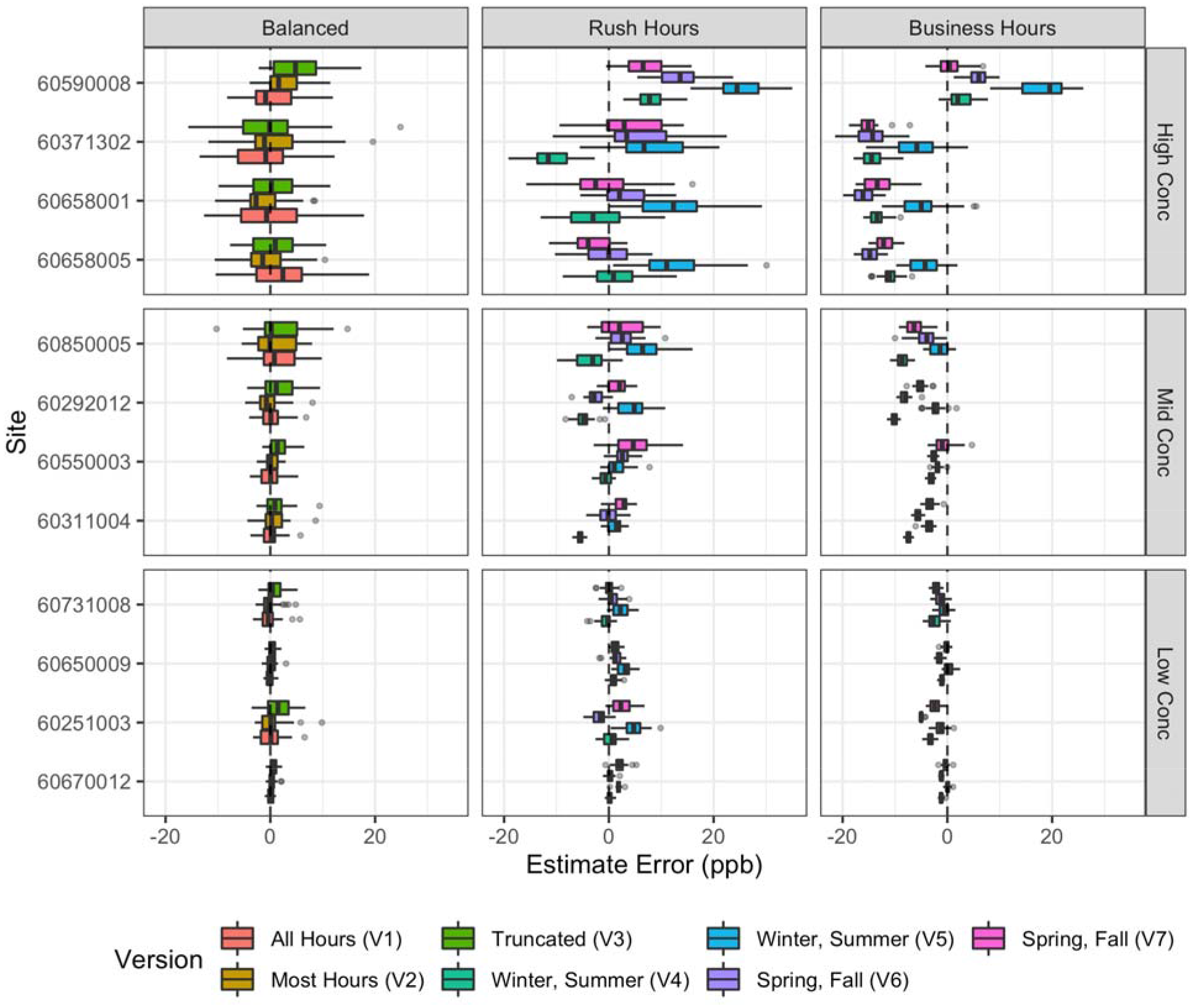
Site-specific NOx measurement error for short-term designs (N = 30 campaigns) as compared to the true annual average at that site (long-term Balanced Design Version 1). Showing a stratified random sample of 12 sites, stratified by whether their true concentration was in the low (<25^th^ percentile), middle (25^th^ −75^th^ percentile) or high (>75^th^ percentile) concentration category and arranged within each stratum with lower concentration sites being closer to the bottom.

### 3.3 Model Predictions

The PLS model of the true annual average had a root mean square error (RMSE) of 7.2 ppb and a mean square error-based coefficient of determination (R^2^_MSE_) of 0.46.

We compared PLS model predictions from each short-term design to the gold standard model predictions. SI Figure S9 shows the relative standard deviations of predictions by design version, with 1 indicating that design predictions have the same standard deviation as the gold standard model predictions. Overall, the Balanced Design predictions have similar variability to those of the gold standard (range: 0.87-1.28), the Rush Hours Design predictions are more variable (range: 0.90-1.74), and the Business Hours Design predictions are mixed: some less and some more variable (range: 0.73-1.54). Figure 4 displays these comparisons as scatterplots and best fit lines. The scatterplots show that there are a few sites, some of which have high leverage, that have variable predictions in all designs. From the best fit lines, we observe that all short-term Balanced Design versions resulted in the most accurate predictions on average, as indicated by their overlapping general trends along the one-to-one line. The Rush Hours Design versions were more likely to have a positive general trend, while the Business Hours Design versions were more likely to have a negative general trend, indicating, for example, that higher concentrations were more likely to be over- or under-estimated, respectively. However, there was heterogeneity in this overall pattern across the Rush and Business Hours Design versions. Furthermore, there was additional heterogeneity across individual campaigns. The SI contains comparable figures comparing design predictions to the gold standard and additional figures for NO and NO_2_ (SI Figures S10-S13).

**Figure 4.**
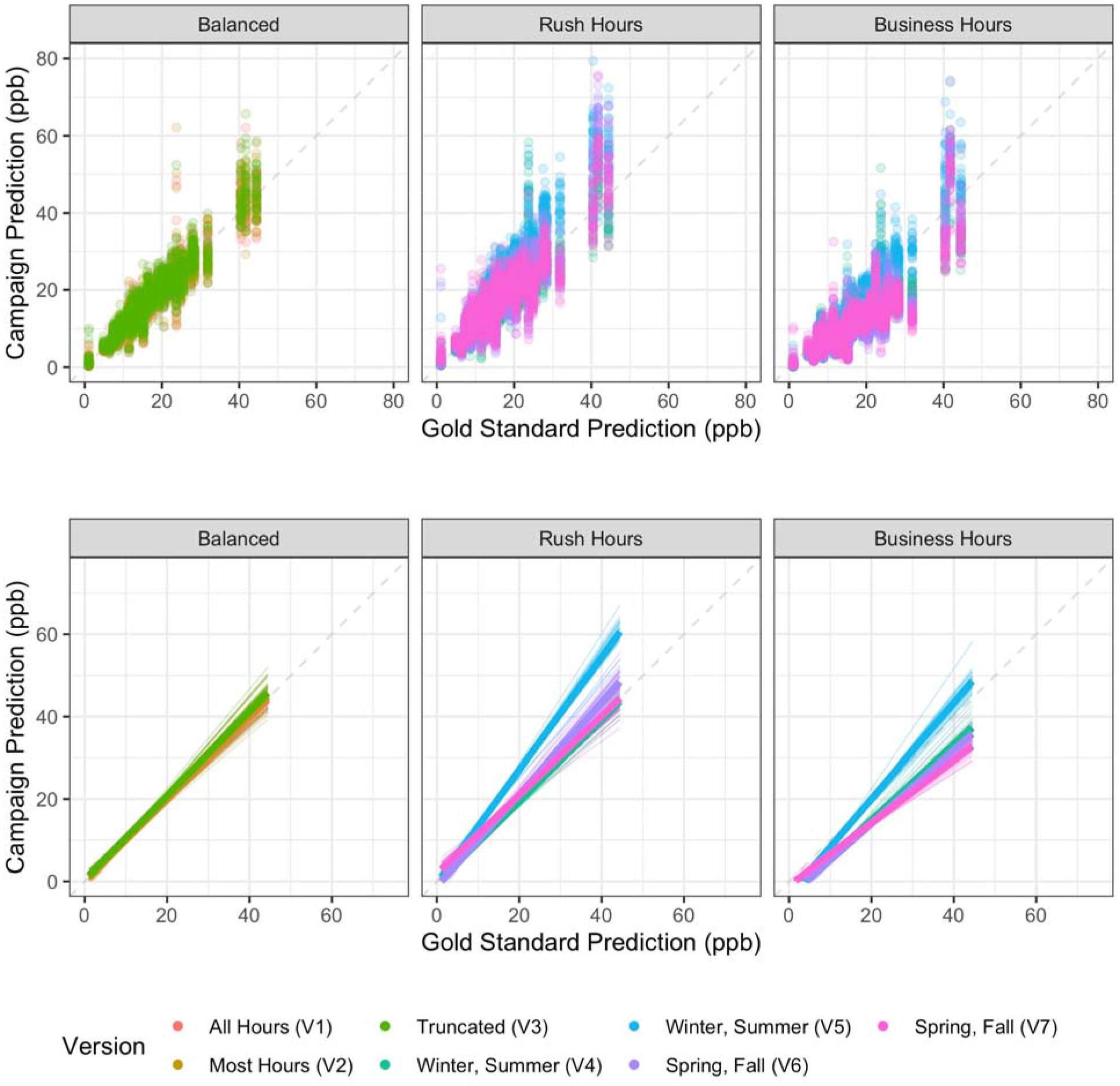
Scatterplots and best fit lines of cross-validated short-term predictions for 30 campaigns vs the gold standard predictions for NOx. Thin transparent lines are individual campaigns, colored by design version; thicker lines are the overall version trend. (One prediction is excluded for clarity from the Rush Hours Version 4 scatterplot at x=24 ppb, y=109 ppb [site 60731016] but included in the line plots.)

Figure 5 shows site-specific comparisons of predictions across 30 short-term campaigns relative to the gold standard predictions for a stratified random sample of 12 sites in order to characterize relative bias (see SI Figure S14 for all sites). Overall, the short-term Balanced Design predictions had a median (IQR) bias of 0.2 (−1 – 1.4) ppb relative to the gold standard predictions (see SI Table S7 for details). All Balanced Design predictions were very similar to the gold standard predictions, though some sites frequently had larger biases. The Rush Hours and Business Hours Design versions were more likely to consistently produce biased site predictions, with a median (IQR) bias of 1.2 (−1.2 – 4) ppb and −3.8 (−6.6 – −1.4) ppb, respectively. While the Rush Hours Design versions generally resulted in higher predictions across sites (with some inconsistency across versions for a few sites), the Business Hours Design versions resulted in predictions that were both lower and higher than the gold standard predictions across sites. There were also a few sites that tended to have more biased and/or more variable predictions relative to the gold standard across all designs. We observed similar patterns when looking at estimate (rather than prediction) biases (See Figure 3, SI Figure S8).

**Figure 5.**
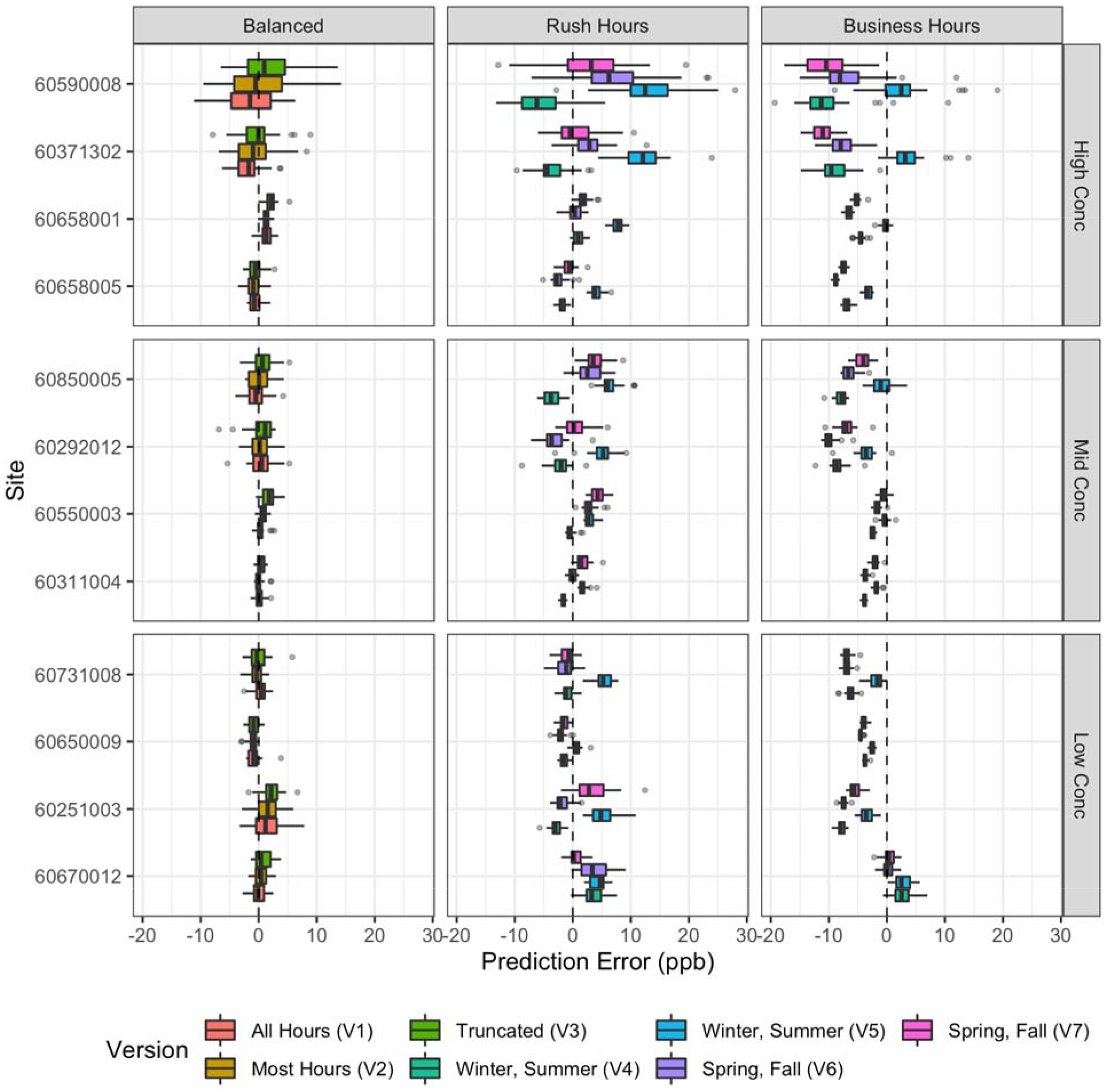
Site-specific NOx prediction errors for short-term designs (N = 30 campaigns) as compared to the gold standard predictions (long-term Balanced Design Version 1). Showing a stratified random sample of 12 sites, stratified by whether true concentrations were in the low (Conc < 0.25), middle (0.25 ≤ Conc ≤ 0.75) or high (Conc > 0.75) concentration quantile and arranged within each stratum with lower concentration sites closer to the bottom.

### 3.4 Model Assessment

Figure 6 shows the out-of-sample prediction performances relative to the observations from the true averages (left column) and the specific design (right column), for both the long-term and short-term approaches. The boxplots quantify the distribution of performance statistics across all 30 short-term campaigns while the squares show the performance for the long-term approach of the same design version. When assessed against the true averages, all the Balanced Design versions generally perform better than either the Rush Hours or Business Hours Design versions with higher CV R^2^_MSE_ and CV R^2^_reg_, and lower CV RMSE estimates. This is particularly apparent for the long-term approach. Furthermore, within design the performance for the long-term approach is better than the majority of the short-term campaigns. There is considerable heterogeneity in performance across the Rush Hours and Business Hours Design versions. In contrast, when assessed against observations from the same design, as would typically be done in practice, the role of sampling design on prediction performance is not as evident. The superior performance of the Balanced Design is not as apparent, and some of the Rush Hours and Business Hours Design versions appear to perform better. There are also a few campaigns that show poor performance, even under the Balanced Design. SI Figure S15-S16 show similar results for NO_2_ and NO, with NO showing more variability and some lower performing statistics. Stratifying by whether sites were considered to have high or low variability (based on hourly standard deviation estimates) showed similar R^2^ and RMSE patterns (data not shown).

**Figure 6.**
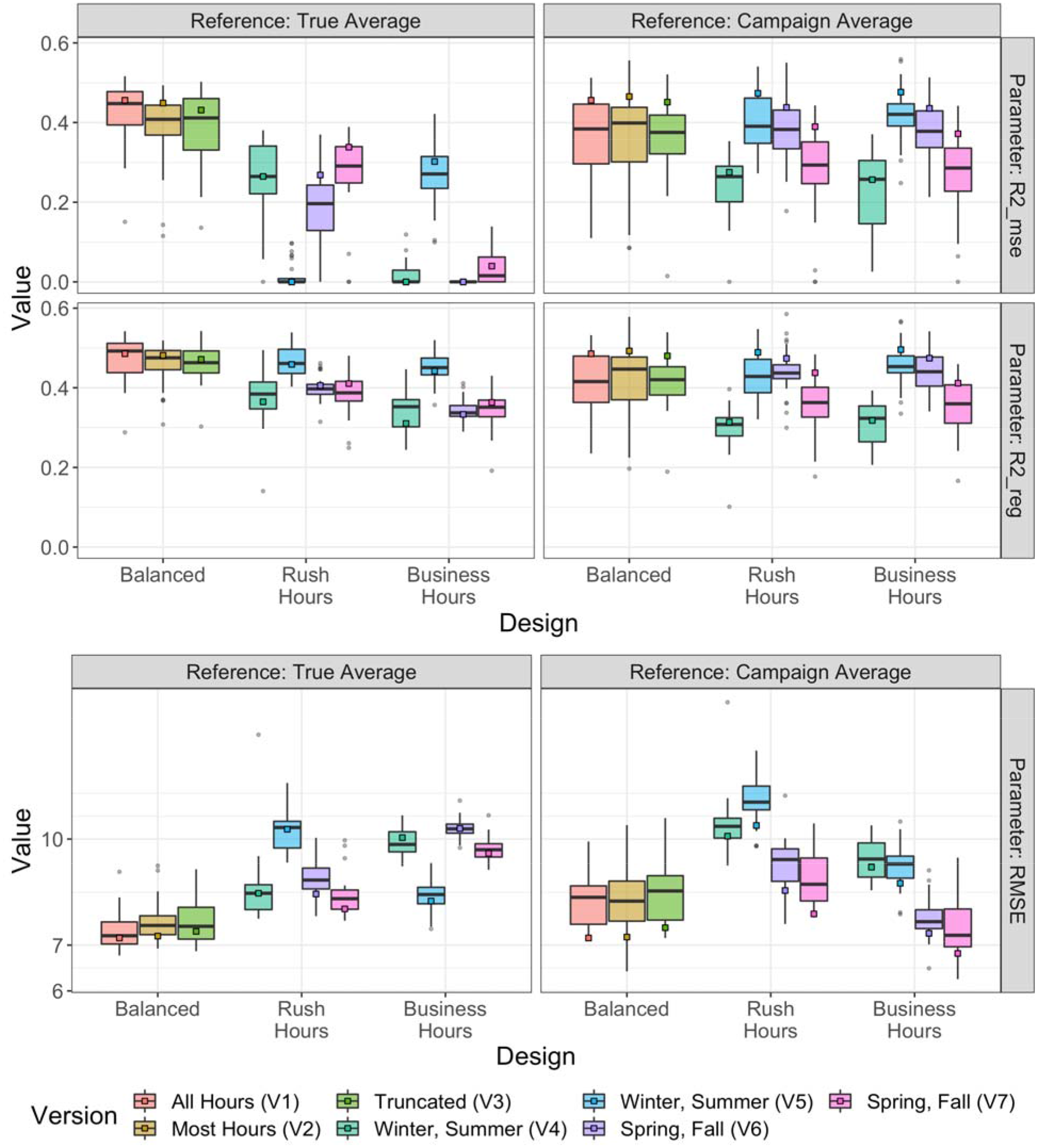
Model performances (MSE-based R2, Regression-based R2, and RMSE), as determined by each campaign’s cross-validated predictions relative to: a) the true averages (long-term Balanced Version 1), and b) its respective campaign averages. Boxplots are for short-term approaches (30 campaigns), while squares are for long-term approaches (1 campaign).

### 3.5 Sensitivity Analyses

Findings were similar for sensitivity analyses (see the SI for NO and NO_2_ results). Figure 7 and

**Figure 7.**
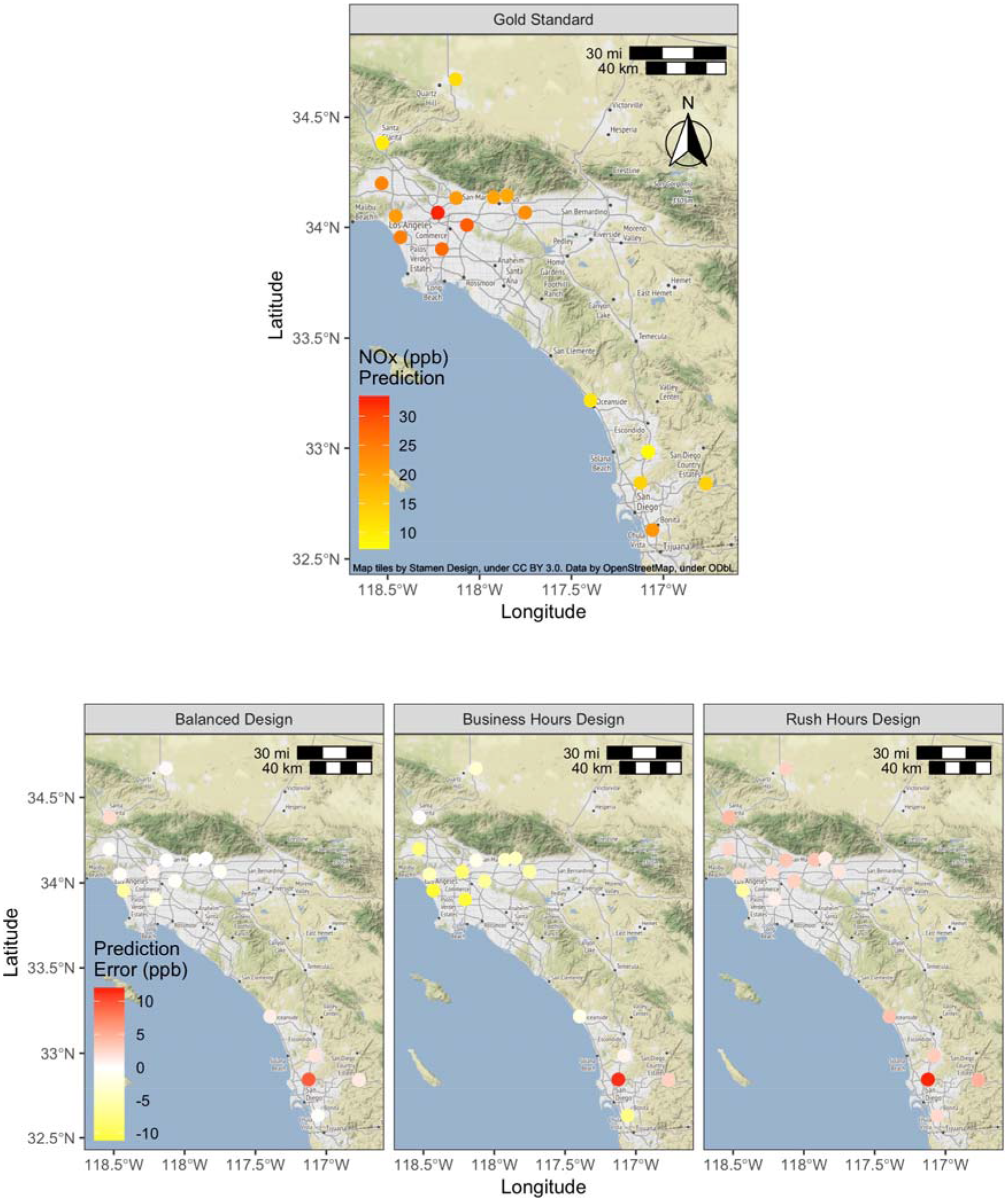
Site predictions from the gold standard campaign (long-term, Balanced Design, All Hours) and prediction errors from each short-term design, as compared to the gold standard campaign, for the Los Angeles-San Diego sensitivity analysis (N = 17 sites).

Table 2 further illustrate the resulting predictions for the Los Angeles-San Diego analysis for the gold standard campaign (long-term Balanced Version) and each of the short-term designs. Short-term designs estimates are for the average site prediction across all simulations and design versions for simplicity. Compared to the gold standard campaign, the median prediction bias (and percent error) for the Balanced, Rush Hours and Business Hours designs was about 0.0 ppb (13.2%), 2.1 ppb (20.4%) and −4.0 (27.5%), respectively.

**Table 2.**
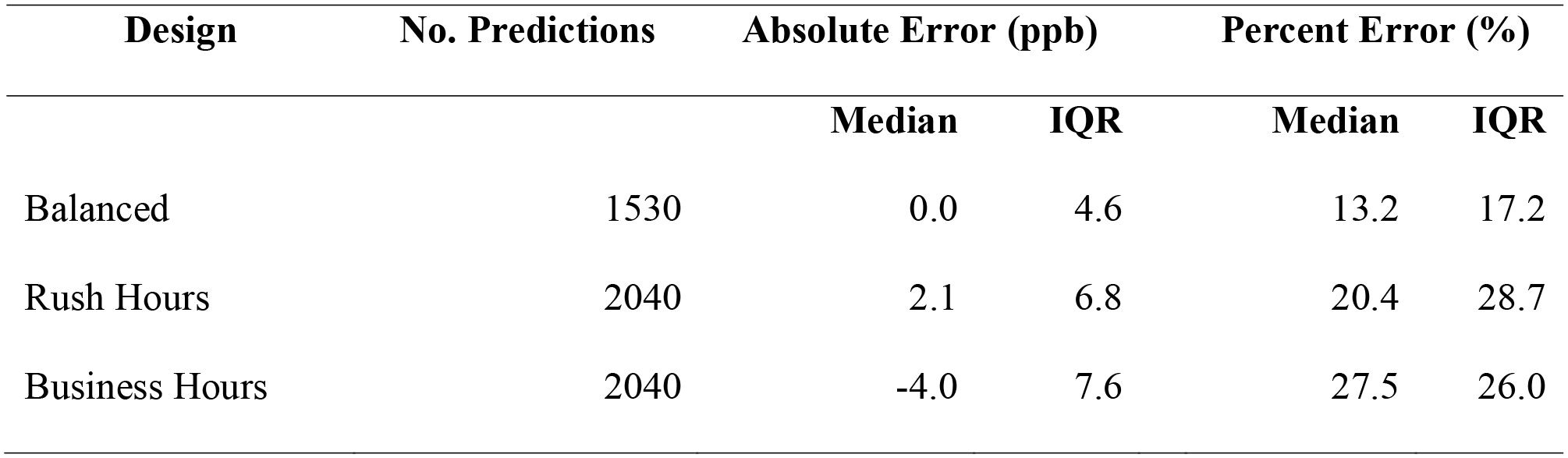
Site prediction error by design relative to the gold standard campaign predictions for the southern California sensitivity analysis (No. Predictions = 17 sites x 30 simulations/version x 3-4 versions/design).

## 4 Discussion

In this paper we have used existing regulatory monitoring data to deepen our understanding of the importance of short-term stationary mobile monitoring study design for application to epidemiologic cohort studies. Others have shown that short-term data can be used to estimate long-term averages.^8, 9^ What has been missing from the literature until now, however, is the impact of short-term stationary mobile monitoring study design on the accuracy and precision of long-term exposure estimates and model predictions, particularly when the goal is to produce predictions for an epidemiologic study. Our results indicate that for designs with a sufficient number of short-term samples at each location (about 28 or more), the design rather than the sampling approach (i.e., sampling duration at a given site) has the largest impact on the estimated annual averages. We focus the rest of this discussion on the short-term approaches for each design, which resemble mobile monitoring, though the long-term approaches produced similar results.

In terms of specific design, we found that all of the Balanced Design versions resulted in similar annual averages as the true averages (long-term Balanced Version 1), while the Rush Hours and Business Hours Design versions were more likely to result in more biased and more or less variable annual average estimates. Specifically, the Rush Hours Design was more likely to overestimate, while the Business Hours Design was more likely to underestimate site averages. This result was likely because the Balanced Design captured much of NOx’s temporal variability by allowing for samples to be collected during each season, day of the week, and all or most times of the day, all periods during which meteorology and traffic activity patterns impact air pollution concentrations (SI Figure S4-S6). The Rush Hours Design, on the other hand, was restricted to two sampling seasons and was more likely to sample during high concentration times of day and days of the week. The Business Hours Design had similar limitations though it was more likely to sample during low concentration times. These conclusions were the same in the Los Angeles-San Diego sensitivity analysis, which is more representative of a geographic area that could be realistically sampled by a mobile campaign.

We found a similar pattern with the predictions: similar predictions across all Balanced Design versions, while most of versions in the Rush Hours tended to overpredict and those in the Business Hours tended to underpredict. However, this varied by design version, suggesting that the particular four weeks of sampling are an important source of heterogeneity in the results. The predictions were more variable for all Rush Hours Design versions and one Business Hours Design version (SI Figure S9). One Business Hours Design version was less variable, while two versions were about the same relative to the gold standard predictions.

The similarity in annual averages and predictions across all of the Balanced Design versions suggests that campaigns with slightly reduced sampling hours (for example, due to logistical constraints) should to a large degree still produce unbiased annual averages at most sites. On the other hand, campaigns that follow more temporally restricted sampling designs such as the Rush Hours and Business Hours Designs may produce systematically biased results, with the degree and direction of error being heavily impacted by the sampling window that happens to be selected.

At the site level, we saw that while any individual study campaign had the potential to produce biased estimates and predictions, the Rush Hours and Business Hours Designs were more likely to do so than the Balanced Design. The direction and magnitude of bias varied by site and depended upon the sampling design and the typical seasonal, day of week, and time of day patterns of pollution at that site. This suggests a simple correction factor to time-adjust short-term measurements based on long-term observations at a small number of reference sites, for example using regulatory fixed sites, is unlikely to fully adjust for bias at the site level.^22^ While many past campaigns have taken this approach to account for the fact that short-term stationary mobile sampling inherently misses some observations, this approach makes a strong assumption that all sites have the same temporal trends. SI Figure S17 – S19 illustrate the temporal trends for sites included in the Los Angeles-San Diego analysis and clearly shows how lower concentration “background” sites are also more likely to have less temporal variation when compared to other sites. Using these “background” sites (or any other site for that matter) to adjust readings at other sites would not substantially reduce the bias from an unbalanced sampling design. This may be especially pertinent for mobile monitoring campaigns since their increased spatial coverage is more likely to capture localized pollution hotspots that may have even more temporal variation. We thus argue that sampling design should be prioritized in mobile monitoring campaigns. Analytical methods such as temporal adjustment factors, on the other hand, should be further investigated to establish their true value given their strong assumptions.

Furthermore, non-balanced designs may misrepresent some sites more than others and lead to differential exposure misclassification in epidemiologic studies since higher concentration sites were more likely to have greater degrees of bias and variation (Figure 4 – Figure 5). Thus, while non-balanced designs may be appropriate for non-epidemiologic purposes including characterizing the spatial impact of traffic related air pollutants during peak hours for urban planning and policy purposes, these could be misleading in epidemiologic applications.

In this study we were able to evaluate prediction model performance against the true annual average NOx exposure as well as against the observations typically available for model performance assessment. Performance assessment against the true averages indicates that the Balanced Design is clearly the best, and that there is little degradation in performance across design versions. This means it is possible to design high quality short-term stationary mobile monitoring studies that accommodate some measure of logistical feasibility, for example, by not requiring sampling in the middle of the night. In contrast, the performance of the Rush Hours and Business Hours Designs is comparatively worse, indicating that the logistically appealing approach that samples only four weeks during two seasons, during daytime hours, and only during weekdays is inadequate for providing high quality estimates of annual averages. Further, the performance of these designs varies considerably and unpredictably depending upon the specific pair of two-week periods that are selected for sampling. Additionally, comparison of the two R^2^ estimates (R^2^_MSE_ and R^2^_reg_) indicates that not all of their poor performance is due to the inability to predict the same value as the truth (R^2^_MSE_), but due to systematic bias in the design. As noted earlier, R^2^_MSE_ assesses whether two measurements are the same - along the 1-1 line, whereas R^2^_reg_ simply assesses whether they are linearly associated. SI Figure S13, for example, shows that Balanced Designs generally produce predictions that are more similar to the “true” estimates from a gold standard campaign (closer to the 1-1 line), whereas the Rush Hours and, in particular, the Business Hours Designs are more likely to produce predictions away from the 1-1 line. This results in the Balanced Designs having R^2^_MSE_ estimates that are only slightly lower than R^2^_reg_ estimates, whereas this drop in performance is greater for the Rush Hours and Business Hours Designs, as seen in Figure 6.

Further, it is notable that the standard approach to model assessment, comparing model predictions to observations collected during the sampling campaign, doesn’t clearly reveal the superior performance of the Balanced Design or the inherent flaws of the Rush Hours and Business Hours Designs. In fact, some of the Rush Hours and Business Hours Design versions perform better than the Balanced Design when evaluated against the campaign’s observations. This is because the evaluation doesn’t take into account that the observations are biased because of the sampling design.

It is notable that the performance of short-term stationary mobile campaigns were fairly consistent with, though generally slightly worse than, the performance observed in the longer-term campaigns for each design version (Figure 6). However, occasionally there was an “unlucky” short-term campaign with meaningfully poorer performance than the other campaigns of the same design. This was more likely in the non-balanced designs, though even the Balanced Design versions had 1-2 of the 30 campaigns (∼3-6%) with notably worse performance as quantified by R^2^. It may be possible that this result is driven by a few high-leverage outlier sites that impact the prediction model performance. In practice, mobile monitoring study investigators are likely to investigate high-leverage sites and address their influence in their prediction modeling.

Our study focused on short-term stationary mobile campaigns with 28 repeat samples per site. We did not consider campaigns with fewer or more visits. As evident in SI Figure S2, the percent error in estimating the annual average from fewer than 25 visits skyrockets, suggesting that site estimates will be considerably noisier in mobile campaigns with few repeat visits, regardless of the study design. Prediction model performance is thus likely to decrease as the number of visits per site decrease. Logistically, it is also difficult to achieve balance in sampling over time across season, day of week, and time of day with fewer than 28 samples per site. Furthermore, we note that this study focused on a few generalizable, common designs in the literature, though many other approaches have been taken. We expect that the variety of mobile campaign designs that have been implemented will all produce slightly different results.

In putting these results in context, it is important to recognize that in this simulation study we are using NOx hourly averages to approximate much shorter-term sampling durations (e.g., a few minutes or less) than would be collected during a mobile monitoring campaign. Shorter duration sampling will affect the noise in the data, to an amount that depends on the environment (e.g., temporal patterns in the concentrations of the pollutant being measured) and the instruments. For comparison, however, our additional evaluations of minute-level data suggest that the decrease in percent error in going from two-minute to hour-long samples is at most a few percent because of serial correlation in the data. This thus gives us confidence that the findings from this work are still generalizable to more common, shorter-term stationary monitoring campaigns with sampling periods closer to a few minutes.

Further, our study took place throughout California, a large, geographically diverse area with varying climate profiles.^36^ While such a large sampling domain would be challenging for a real-world mobile monitoring campaign, the overall conclusions of this study – the importance of temporally-balanced sampling, are also supported in the Los Angeles-San Diego sensitivity analysis. In terms of the siting criteria for the regulatory monitoring sites where the data came from, locations are generally meant to capture representative population exposures, including near roadway, at various spatial scales ranging from microscales (< 100 m range) to regional scales in order to inform regulatory compliance.^37, 38^ This should thus have provided us with decent spatial coverage and concentration variability. Most air pollution studies, in fact, rely on this network of regulatory monitors.^39^ Still, when compared to most mobile monitoring campaigns, this study’s larger domain and reduced exposure variability may have produced lower prediction model performances than would be expected from mobile monitoring campaigns.

Another distinction is that while we sampled measurements within sites at random, mobile campaigns typically sample from sites along a fixed route or in a designated area. The actual sampling scheme will thus depend on the exact route developed and the number of platforms deployed, both of which are beyond the scope of this paper. In general, sampling along a route also induces some spatial correlation in the mobile monitoring data. This dependence is often overlooked in mobile monitoring campaigns and was not addressed in this study. Furthermore, we did not consider the importance of the distribution of sampling locations in this study, which is particularly relevant when the exposure assessment goal is an epidemiologic application. Selecting sites that are representative of the target cohort’s residence locations will ensure the spatial compatibility assumption is met, which is an important way to reduce the role of exposure measurement error in epidemiologic inference.^40^ This consideration is especially relevant for mobile monitoring near major sources (e.g., airports, marine activity, and industry),^8, 9, 41–47^ which may or may not represent a study cohort of interest.

Our evaluation focused on NOx, NO, and NO_2_, which are quickly and moderately decaying air pollutants (concentrations reach background levels approximately 400-600 m from roadway sources).^21^ Campaigns that measure these pollutants may be more susceptible to sampling design than campaigns that measure less spatially- and/or temporally-variable pollutants such as PM_2.5_.^27^ We selected NOx, NO, and NO_2_ because these traffic-related pollutants are often measured in short-term campaigns, and data for these pollutants are more widely available. Non-criteria pollutants, for example ultrafine particulates (UFP), however, have also received increasing attention in recent years given their emerging link to adverse health effects.^48–51^ Still, high-quality information about their spatial distribution is essentially absent, and most studies have implemented short-term mobile sampling approaches^47^ that may not be temporally balanced and potentially be misleading. Finally, while other discrepancies surely exist between this simulation study and realized mobile monitoring campaigns, we expect our overall conclusions on the importance of temporally-balanced sampling to be remain the same.

An important next step in this work is to understand whether the differences in exposure estimates that we observed across study designs have a meaningful impact on epidemiologic inferences. This is of particular interest considering that year-around, balanced designs are resource-intensive and rare, while shorter, more convenient campaigns are more common in the literature. More research is needed to better understand how and whether unbalanced mobile monitoring campaigns may contribute high quality exposure assessments for epidemiology. Regardless of design, we expect that the predictions from all of the campaigns will result in both classical-like and Berkson-like error.^40, 52–54^ Specifically, the predictions capture only part of the true long-term exposure (Berkson-like error), while the parameters in the prediction model are inherently noisy (classical-like error). However, these measurement error methods have not to date considered exposure assessment study design, beyond considering the importance of spatial compatibility, i.e., that distribution of monitoring locations is the same as the distribution of participant locations. Our work suggests that deeper understanding of the role of exposure assessment design on epidemiologic inference is an important area of research.

### 4.1 Conclusions and Recommendations for Mobile Monitoring Campaigns

Mobile monitoring study design should be an important consideration for campaigns aiming to assess long-term exposure in an epidemiologic cohort. Given the temporal trends in air pollution, campaigns should implement balanced designs that sample during all seasons of the year, days of the week, and hours of the day in order to produce unbiased annual averages. Nonetheless, restricting the sampling hours in balanced designs, for example due to logistical considerations, will still generally produce unbiased estimates at most sites. On the other hand, unbalanced sampling designs like those often seen in the literature are more likely to produce biased annual estimates, with some sites being more biased than others. And while predictions from these restricted designs may at times perform similarly to balanced designs (or, more problematically, may erroneously *appear* to perform similarly when evaluated against measurements which are themselves biased samples), this performance may strongly depend on the exact sampling period chosen and may thus be difficult or impossible to anticipate prior to conducting a new sampling campaign. Furthermore, the differential exposure misclassification that may result from these designs may be problematic in epidemiologic investigations. Finally, studies that implement unbalanced sampling designs are likely to have hidden exposure misclassification given that both the observations and model predictions may be systematically incorrect. By implementing a balanced sampling design, campaigns can thus increase their likelihood of capturing accurate annual average exposure averages.

## Data Availability

Air pollution data are available through the EPA. The covariates used in this analysis for regulatory sites are freely available through various online sources and may be available from the authors upon request.

https://aqs.epa.gov/aqsweb/airdata/download_files.html

