## Supplemental Information for "Design and evaluation of mobile monitoring campaigns for air pollution exposure assessment in epidemiologic cohorts"

Figure S16. NO Model performances ($R^2_{MSE}$, $R^2_{REG}$, and RMSE), as determined by each campaign's cross-validated predictions relative to: a) the true averages (long-term Balanced Version 1), and b) its respective campaign averages. Boxplots are for short-term approaches (30 campaigns),while squares are for long-term approaches (1 campaign). A few influential outliers influenced these performance statistics more so than for NOx and $NO_2$. .. 28

#### List of Equations

### 1 Methods

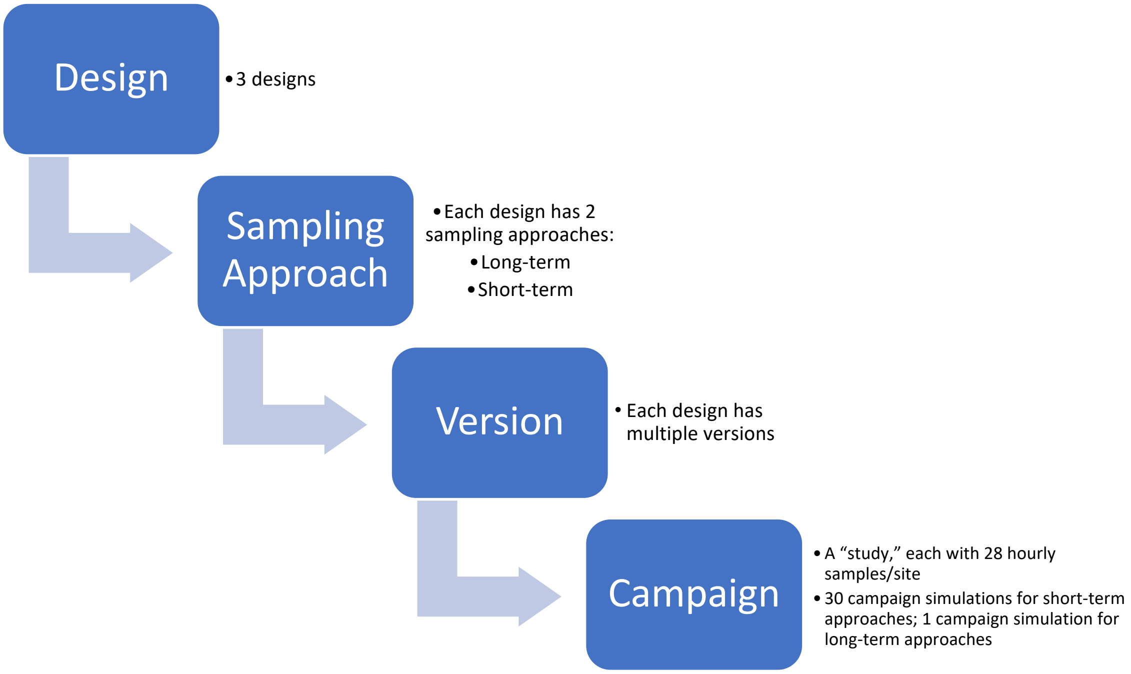

*Figure S1. Hierarchical structure of simulations.*

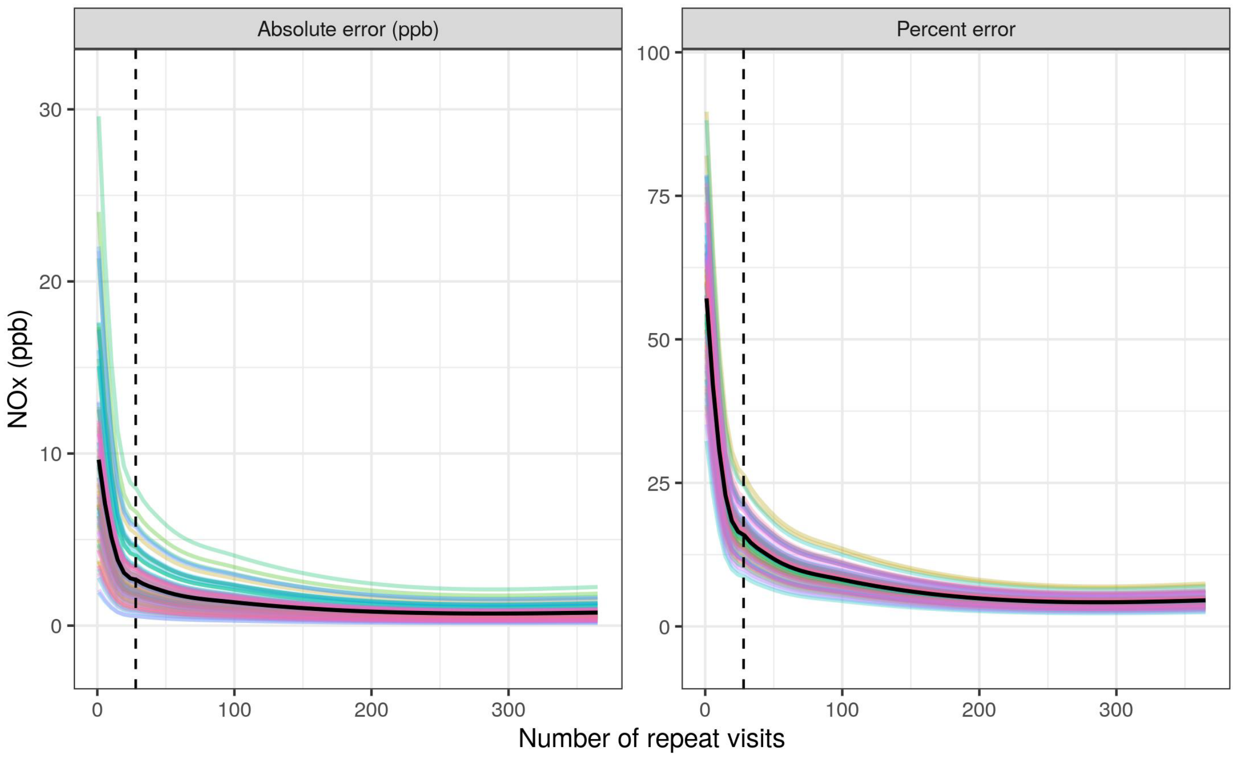

*Figure S2. Loess lines for absolute and percent error of the NOx annual average (ppb), averaged across 10,000 random samples and 69 sites, by number of repeat visits. The colored curves are for individual sites, the black curve is the overall trend, and the dashed vertical line is for 28 repeat visits.*

*Table S1. Two-week sampling windows for the Rush Hours and Business Hours designs*[a]

| Version | Season | Start | End |
|---|---|---|---|
| 4 | summer | 2016-06-20 | 2016-07-03 |
| 4 | winter | 2016-02-27 | 2016-03-11 |
| 5 | summer | 2016-08-07 | 2016-08-20 |
| 5 | winter | 2016-01-15 | 2016-01-28 |
| 6 | spring | 2016-04-15 | 2016-04-28 |
| 6 | fall | 2016-09-25 | 2016-10-08 |
| 7 | spring | 2016-05-15 | 2016-05-28 |
| 7 | fall | 2016-11-24 | 2016-12-07 |

[a] the same two-week periods for each version were used for all sites

*Table S2. Geocovariates and buffers included in PLS regression (n = 321)*

| Kind | Covariate | Buffers | Description |
|---|---|---|---|
| airports | log_m_to_airp | | log meters to closest airport |
| airports | log_m_to_l_airp | | log meters to closest large airport |
| bus | log_m_to_bus | | log meters to closest bus route |
| coast | log_m_to_coast | | log meters to closest coastline |
| commercial and services | log_m_to_comm | | log meters to closest commercial and services area |
| commercial and services | lu_comm_p | 50, 100, 150, 300, 400, 500, 750, 1000, 1500, 3000, 5000, 10000, 15000 | proportion of commercial land use |
| elevation | elev_above | 1000, 5000 | number of points (out of 24) more than 20 m and 50 m uphill of a |

| | | | |
|---|---|---|---|
| | | | location for a 1000 m and 5000 m buffer, respectively |
| elevation | elev_at_elev | 1000, 5000 | number of points (out of 24) within 20 m and 50 m of the location' elevation for a 1000 m and 5000 m buffer, respectively |
| elevation | elev_below | 1000, 5000 | number of points (out of 24) more than 20 m and 50 m downhill of a location for a 1000 m and 5000 m buffer, respectively |
| elevation | elev_elevation | | elevation above sea level in meters |
| emissions/air pollutants | em_CO_s | 3000, 15000, 30000 | sum of major CO emissions from stacks |
| emissions/air pollutants | em_NOx_s | 3000, 15000, 30000 | sum of major NOx emissions from stacks |
| emissions/air pollutants | em_PM10_s | 3000, 15000, 30000 | sum of major PM10 emissions from stacks |
| emissions/air pollutants | em_PM25_s | 3000, 15000, 30000 | sum of major PM2.5 emissions from stacks |
| emissions/air pollutants | em_SO2_s | 3000, 15000, 30000 | sum of major SO2 emissions from stacks |
| emissions/air pollutants | no2_behr_2005 | | Columnar NO2 for 2005 |
| emissions/air pollutants | no2_behr_2006 | | Columnar NO2 for 2006 |
| emissions/air pollutants | no2_behr_2007 | | Columnar NO2 for 2007 |
| imperviousness | imp_a | 50, 100, 150, 300, 400, 500, 750, 1000, 3000, 5000 | average imperviousness |

| | | | |
|---|---|---|---|
| land use | lu_bays_p | 3000, 5000, 10000, 15000 | proportion of land with bays and estuaries |
| land use | lu_crop_p | 100, 150, 300, 400, 500, 750, 1000, 1500, 3000, 5000, 10000, 15000 | proportion of cultivated crops such as orchards, vineyards, grains |
| land use | lu_green_p | 750, 1000, 1500, 3000, 5000, 10000, 15000 | proportion of evergreen forest land |
| land use | lu_grove_p | 750, 1000, 1500, 3000, 5000, 10000, 15000 | proportion of orchards, groves, vineyards, nurseries |
| land use | lu_herb_range_p | 1000, 1500, 3000, 5000, 10000, 15000 | proportion of herbaceous rangeland |
| land use | lu_industrial_p | 150, 300, 400, 500, 750, 1000, 1500, 3000, 5000, 15000 | proportion of industrial land use |
| land use | lu_mine_p | 3000, 5000, 10000 | proportion of land with strip mines, quarries, and gravel pits |
| land use | lu_mix_forest_p | 10000, 15000 | proportion of mixed forest land |
| land use | lu_mix_range_p | 1500, 3000, 5000, 10000 | proportion of mixed rangeland |
| land use | lu_mix_urban_p | 150, 300, 400, 500, 750, 1000, 1500 | proportion of mixed urban or built-up land |
| land use | lu_oth_urban_p | 400, 500, 750, 1000, 1500, 5000 | proportion of other urban or built-up land |
| land use | lu_reservior_p | 5000 | proportion of land with reservoiurs |
| land use | lu_resi_p | 50, 100, 150, 300, 400, 500, 750, 1000, 1500, 3000, 5000, 10000, 15000 | Proportion of residential land use |

| | | | |
|---|---|---|---|
| land use | lu_shrub_p | 400, 500, 750, 1000, 1500, 3000, 5000, 10000, 15000 | proportion of shrubland |
| land use | lu_transition_p | 750, 1000, 1500 | proportion of transitional land use |
| land use | lu_unspec_p | 10000, 15000 | proportion of unspecified land use |
| land use | rlu_barren_p | 3000, 5000 | proportion of barren land |
| land use | rlu_crop_p | 750, 1000, 3000, 5000 | proportion of cropland and pasture land |
| land use | rlu_dev_hi_p | 50, 100, 150, 300, 400, 500, 750, 1000, 3000, 5000 | proportion of highly developed land (e.g., commercial and services; industrial; transportation, communication and utilities) |
| land use | rlu_dev_lo_p | 50, 100, 150, 300, 400, 500, 750, 1000, 3000, 5000 | proportion of low developed land (e.g., residential) |
| land use | rlu_dev_med_p | 50, 100, 150, 300, 400, 500, 750, 1000, 3000, 5000 | proportion of medium developed land (e.g., residential) |
| land use | rlu_dev_open_p | 50, 100, 150, 300, 400, 500, 750, 1000, 3000, 5000 | proportion of developed open land |
| land use | rlu_grass_p | 50, 100, 150, 300, 400, 500, 750, 1000, 3000, 5000 | proportion of grasslands, herbaceous vegetation |
| land use | rlu_herb_wetland_p | 5000 | proportion of herb (nonforested) wetland |
| land use | rlu_mix_forest_p | 3000, 5000 | proportion of mixed forest |
| land use | rlu_pasture_p | 1000, 3000, 5000 | proportion of pasture, hay land |
| land use | rlu_shrub_p | 400, 500, 750, 1000, 3000, 5000 | proportion of shrubland |

| | | | |
|---|---|---|---|
| NDVI | ndvi_q25_a | 250, 500, 1000, 2500, 5000, 7500, 10000 | NDVI (25th quantile) |
| NDVI | ndvi_q50_a | 250, 500, 1000, 2500, 5000, 7500, 10000 | NDVI (50th quantile) |
| NDVI | ndvi_q75_a | 250, 500, 1000, 2500, 5000, 7500, 10000 | NDVI (75th quantile) |
| NDVI | ndvi_summer_a | 250, 500, 1000, 2500, 5000, 7500, 10000 | average summer time NDVI |
| NDVI | ndvi_winter_a | 250, 500, 1000, 2500, 5000, 7500, 10000 | average winter time NDVI |
| population | pop_s | 500, 1000, 1500, 2000, 2500, 3000, 5000, 10000, 15000 | 2000 population density |
| port | log_m_to_s_port | | log meters to closest small port |
| port | lu_transport_p | 300, 400, 500, 750, 1000, 1500, 3000, 5000 | proportion of transportation, communications, and utilities land |
| railroads, rail yards | log_m_to_rr | | log meters to closest railroad |
| railroads, rail yards | log_m_to_ry | | log meters to closest rail yard |
| roads | intersect_a1_a1_s | 3000 | intersect_a1_a1_s |
| roads | intersect_a1_a2_s | 3000 | intersect_a1_a2_s |
| roads | intersect_a1_a3_s | 1000, 3000 | number of a1-a3 road intersections |
| roads | intersect_a2_a2_s | 3000 | number of a2-a2 road intersections |
| roads | intersect_a2_a3_s | 3000 | number of a2-a3 road intersections |

| | | | |
|---|---|---|---|
| roads | intersect_a3_a3_s | 500, 1000, 3000 | number of a3-a3 road intersections |
| roads | ll_a1_s | 500, 750, 1000, 1500, 3000, 5000 | length of a1 roads |
| roads | ll_a2_s | 1500, 3000, 5000 | length of a2 roads |
| roads | ll_a3_s | 50, 100, 150, 300, 400, 500, 750, 1000, 1500, 3000, 5000 | length of a3 roads |
| roads | log_m_to_a1 | | log meters to closest a1 road |
| roads | log_m_to_a1_a1_intersect | | log meters to closest a1-a1 road intersection |
| roads | log_m_to_a1_a2_intersect | | log_m_to_a1_a2_intersect |
| roads | log_m_to_a1_a3_intersect | | log meters to closest a1-a3 road intersection |
| roads | log_m_to_a2 | | log meters to closest a2 road |
| roads | log_m_to_a2_a2_intersect | | log meters to closest a2-a2 road intersection |
| roads | log_m_to_a2_a3_intersect | | log meters to closest a2-a3 road intersection |
| roads | log_m_to_a3 | | log meters to closest a3 road |
| roads | log_m_to_a3_a3_intersect | | log meters to closest a3-a3 road intersection |
| truck routes | log_m_to_truck | | log meters to closest truck route |
| truck routes | tl_s | 750, 1000, 1500, 3000, 5000, 10000, 15000 | length of truck routes |
| water | log_m_to_waterway | | log meters to closest waterway |
| water | rlu_water_p | 3000, 5000 | proportion of water |

$$MSE_{ref} = \frac{1}{n}\sum_{i=1}^{n}\left(y_{i,ref} - \hat{y}_{i,campaign}\right)^2$$

*Equation S1. Mean squared error (MSE) definition. Where $\hat{y}_{i,campaign}$ is the prediction from a campaign for a given design version; $y_{i,ref}$ is the reference value, either the true annual average or the estimated annual average from the same campaign (the typical approach in practice); and $n$ is the total number of sites.*

$$RMSE_{ref} = \sqrt{MSE_{ref}}$$

*Equation S2. Root mean squared error (RMSE) definition*

$$R_{MSE}^2 = \max\left(0, 1 - \frac{MSE_{ref}}{\frac{1}{n}\sum_{i=1}^{n}\left(y_{i,ref} - \bar{y}_{campaign}\right)^2}\right)$$

*Equation S3. MSE-based $R^2$ ($R_{MSE}^2$) definition. Where $\bar{y}_{campaign}$ is the average across all n sites for a given campaign.*

*Note S1. R packages used in analyses*

dplyr (1.0.6),[1] forcats (0.5.0),[2] ggmap (3.0.0),[3] ggplot2 (3.3.3),[4] ggpubr (0.2.5),[5] ggrepel (0.8.1),[6] ggspatial (1.1.4),[7] glmnet (3.0-2),[8] kableExtra (1.1.0),[9] lubridate (1.7.10),[10] magrittr (1.5),[11] Matrix (1.2-18),[12] modelr (0.1.6),[13] pls (2.7-2),[14] purrr (0.3.3),[15] readr (1.3.1),[16] sf (0.9-5),[17] stringr (1.4.0),[18] tibble (3.1.2),[19] tidyr (1.0.2),[20] tidyverse (1.3.0),[21] VCA (1.4.2)[22]

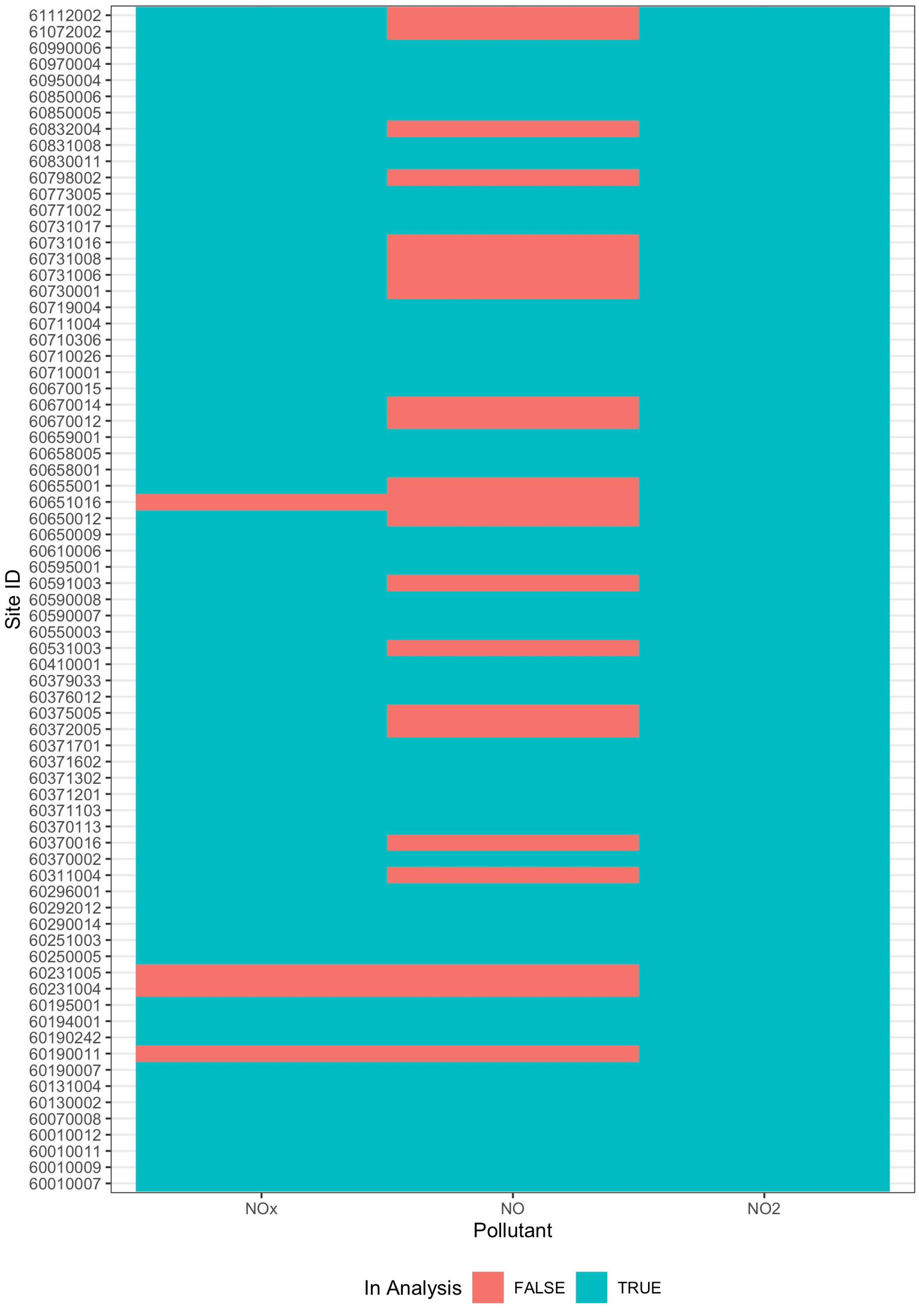

*Figure S3. AQS sites included in the analysis of each pollutant (N=69 NOx, 51 NO, 73 $NO_2$). Site ID is a compilation of the CA state ID (6, the first digit), county ID (next 3 digits), and AQS site ID (last 4 digits).*

#### 2 Hourly Readings

*Table S3. Distribution of the number of hourly and day equivalent (24 samples/day) observations per site[1]*

| Parameter Name | Count | N | Min | Mean | SD | Median | IQR | Max |
|---|---|---|---|---|---|---|---|---|
| Oxides of nitrogen (NOx) | Day Equivalent | 69 | 285 | 337 | 15 | 343 | 17 | 355 |
| Oxides of nitrogen (NOx) | Hours | 69 | 6,836 | 8,090 | 361 | 8,236 | 408 | 8,510 |
| Nitric oxide (NO) | Day Equivalent | 51 | 294 | 338 | 14 | 342 | 14 | 355 |
| Nitric oxide (NO) | Hours | 51 | 7,060 | 8,119 | 339 | 8,216 | 346 | 8,510 |
| Nitrogen dioxide (NO2) | Day Equivalent | 73 | 284 | 337 | 15 | 343 | 17 | 355 |
| Nitrogen dioxide (NO2) | Hours | 73 | 6,825 | 8,077 | 363 | 8,231 | 408 | 8,510 |

[1] N = number of sites.

*Table S4. Distribution of hourly concentrations (ppb)[1]*

| Parameter Name | N | Min | Mean | SD | Median | IQR | Max |
|---|---|---|---|---|---|---|---|
| Oxides of nitrogen (NOx) | 558,207 | -5 | 16 | 21 | 9 | 16 | 427 |
| Nitric oxide (NO) | 414,046 | -5 | 9 | 16 | 4 | 5 | 381 |
| Nitrogen dioxide (NO2) | 589,625 | -3 | 10 | 10 | 7 | 12 | 97 |

[1] N = total number of hourly readings.

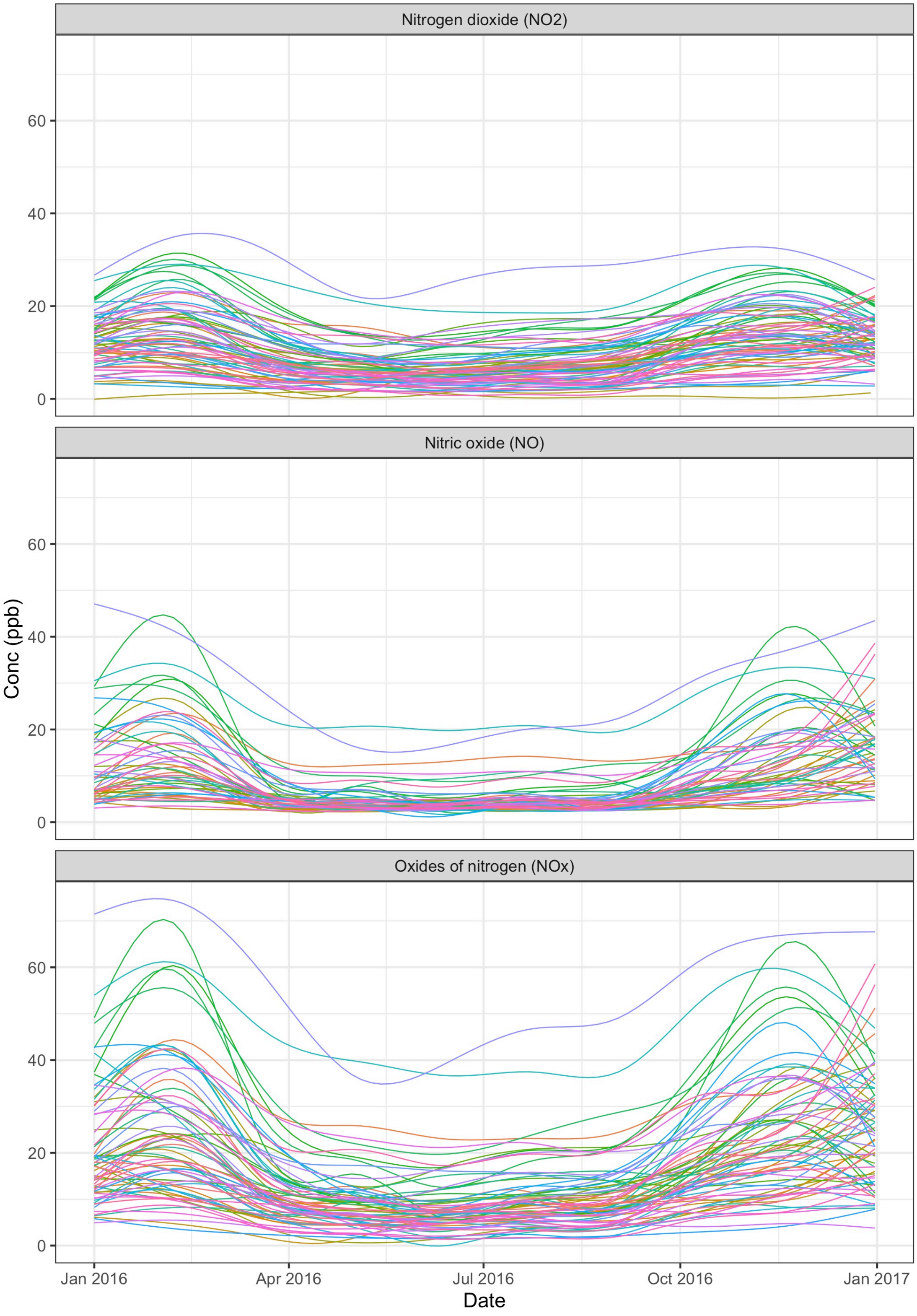

*Figure S4. Concentration trends for NOx, NO, and NO2 over the course of 2016 at AQS sites included in this study (N=69 NOx, 51 NO, 73 $NO_2$). Colored lines are individual sites.*

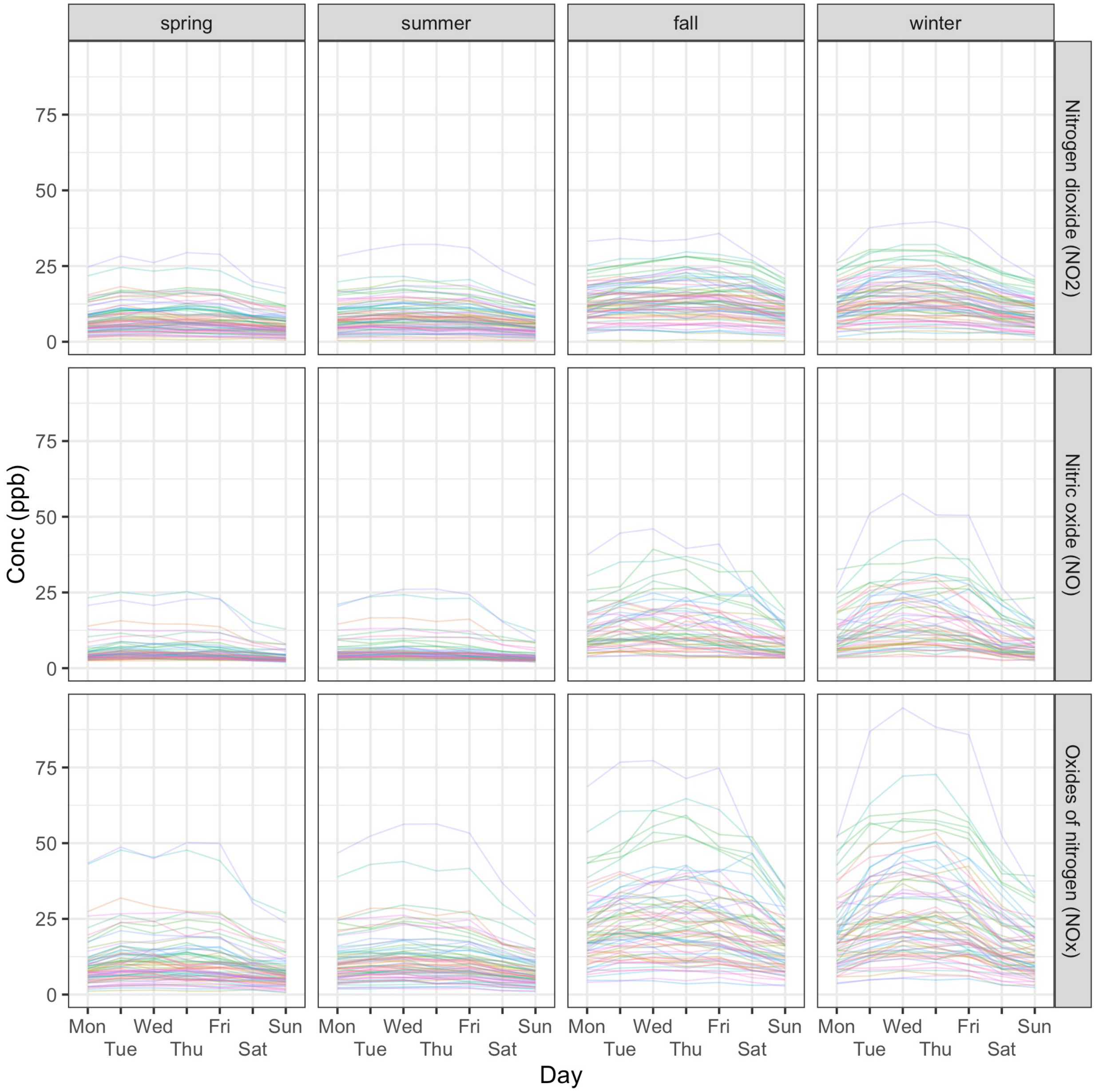

*Figure S5. Concentration trends for NOx, NO, and NO2 by day and season at AQS sites included in this study (N=69 NOx, 51 NO, 73 $NO_2$). Colored lines are individual sites.*

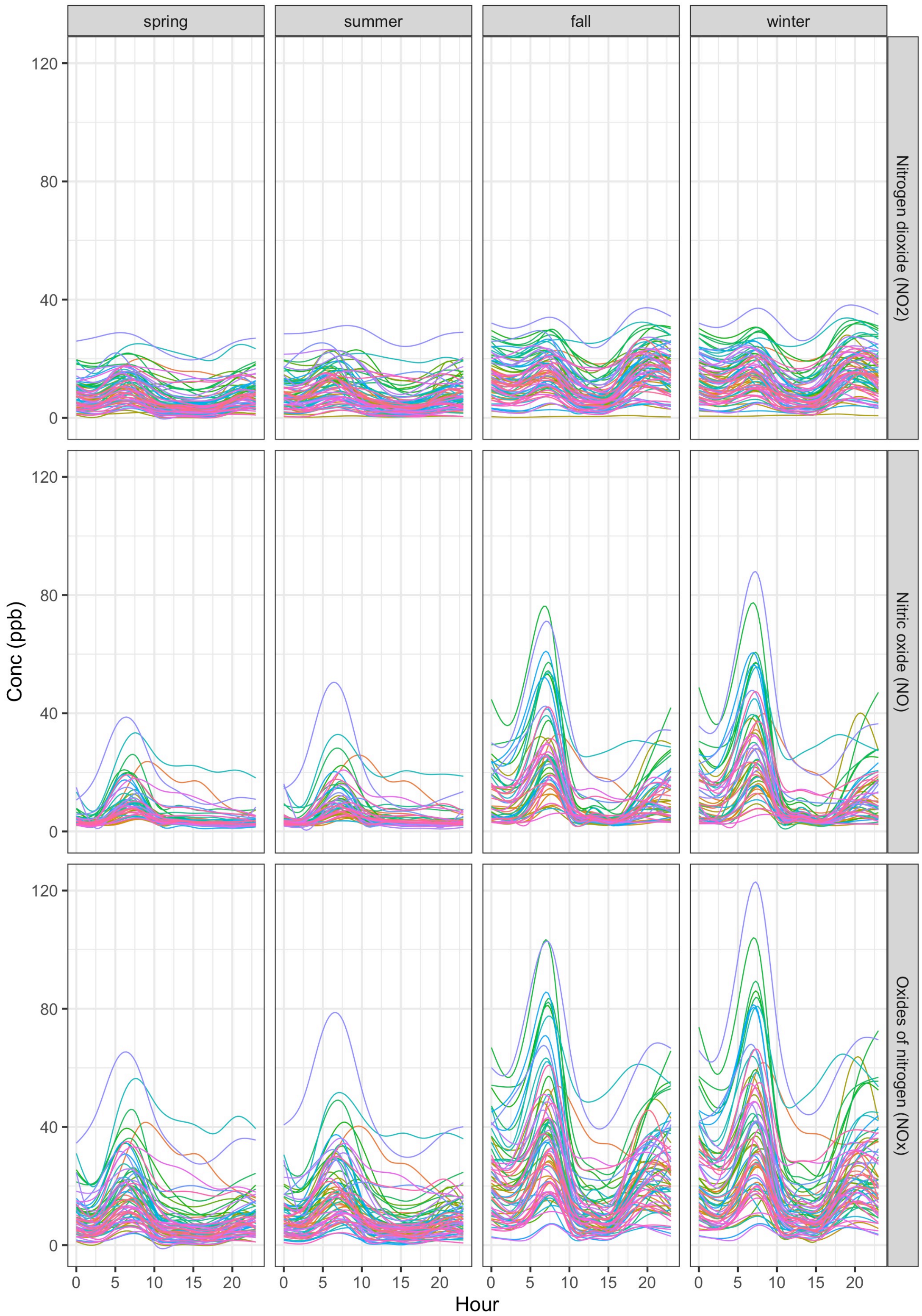

*Figure S6. Concentration trends for NOx, NO, and NO2 by hour and season at AQS sites included in this study (N=69 NOx, 51 NO, 73 $NO_2$). Colored lines are individual sites.*

### 3 Annual Average Estimates

*Table S5. Distribution of annual average NOx estimates from various sampling approaches.*[1]

| Design | Version | Type | N | Min | Q25 | Q50 | Q75 | Max | SD |
|---|---|---|---|---|---|---|---|---|---|
| Balanced | All Hours (V1) | Long-Term | 69 | 3.0 | 9.8 | 14.2 | 20.8 | 55.6 | 9.8 |
| Balanced | All Hours (V1) | Short-Term | 2070 | 1.9 | 9.2 | 13.7 | 21.2 | 69.5 | 10.3 |
| Balanced | Most Hours (V2) | Long-Term | 69 | 3.2 | 9.9 | 14.1 | 20.7 | 55.4 | 9.9 |
| Balanced | Most Hours (V2) | Short-Term | 2070 | 2.1 | 9.4 | 13.9 | 20.9 | 70.8 | 10.3 |
| Balanced | Truncated (V3) | Long-Term | 69 | 3.4 | 10.4 | 14.4 | 21.0 | 55.7 | 10.1 |
| Balanced | Truncated (V3) | Short-Term | 2070 | 2.1 | 9.8 | 14.7 | 21.4 | 66.9 | 10.6 |
| Rush Hours | Winter, Summer (V4) | Long-Term | 69 | 3.5 | 8.3 | 12.1 | 19.4 | 67.1 | 12.0 |
| Rush Hours | Winter, Summer (V4) | Short-Term | 2070 | 2.2 | 8.2 | 12.2 | 19.8 | 78.2 | 12.0 |
| Rush Hours | Winter, Summer (V5) | Long-Term | 69 | 4.6 | 12.3 | 18.8 | 29.6 | 75.9 | 14.5 |
| Rush Hours | Winter, Summer (V5) | Short-Term | 2070 | 3.1 | 12.0 | 18.1 | 29.0 | 95.7 | 14.8 |
| Rush Hours | Spring, Fall (V6) | Long-Term | 69 | 3.1 | 10.4 | 13.6 | 20.4 | 58.8 | 11.3 |
| Rush Hours | Spring, Fall (V6) | Short-Term | 2070 | 1.7 | 9.4 | 13.7 | 20.7 | 70.8 | 11.7 |
| Rush Hours | Spring, Fall (V7) | Long-Term | 69 | 4.4 | 10.6 | 15.7 | 20.2 | 55.7 | 10.0 |
| Rush Hours | Spring, Fall (V7) | Short-Term | 2070 | 2.3 | 10.4 | 15.3 | 20.9 | 69.5 | 10.4 |

| | | | | | | | | | |
|---|---|---|---|---|---|---|---|---|---|
| Business Hours | Winter, Summer (V4) | Long-Term | 69 | 1.6 | 5.5 | 8.1 | 12.8 | 54.6 | 10.6 |
| Business Hours | Winter, Summer (V4) | Short-Term | 2070 | 1.0 | 5.4 | 8.1 | 14.2 | 62.1 | 10.7 |
| Business Hours | Winter, Summer (V5) | Long-Term | 69 | 3.1 | 8.0 | 11.9 | 20.5 | 65.2 | 12.0 |
| Business Hours | Winter, Summer (V5) | Short-Term | 2070 | 2.0 | 8.1 | 11.8 | 19.9 | 73.0 | 12.0 |
| Business Hours | Spring, Fall (V6) | Long-Term | 69 | 1.6 | 5.4 | 7.9 | 12.2 | 53.3 | 9.8 |
| Business Hours | Spring, Fall (V6) | Short-Term | 2070 | 1.1 | 5.1 | 8.0 | 12.1 | 57.1 | 9.8 |
| Business Hours | Spring, Fall (V7) | Long-Term | 69 | 2.3 | 6.1 | 9.7 | 12.7 | 47.2 | 8.7 |
| Business Hours | Spring, Fall (V7) | Short-Term | 2070 | 0.9 | 6.2 | 9.6 | 13.1 | 53.9 | 8.7 |

[1] N = Total number of sites x number of campaigns.

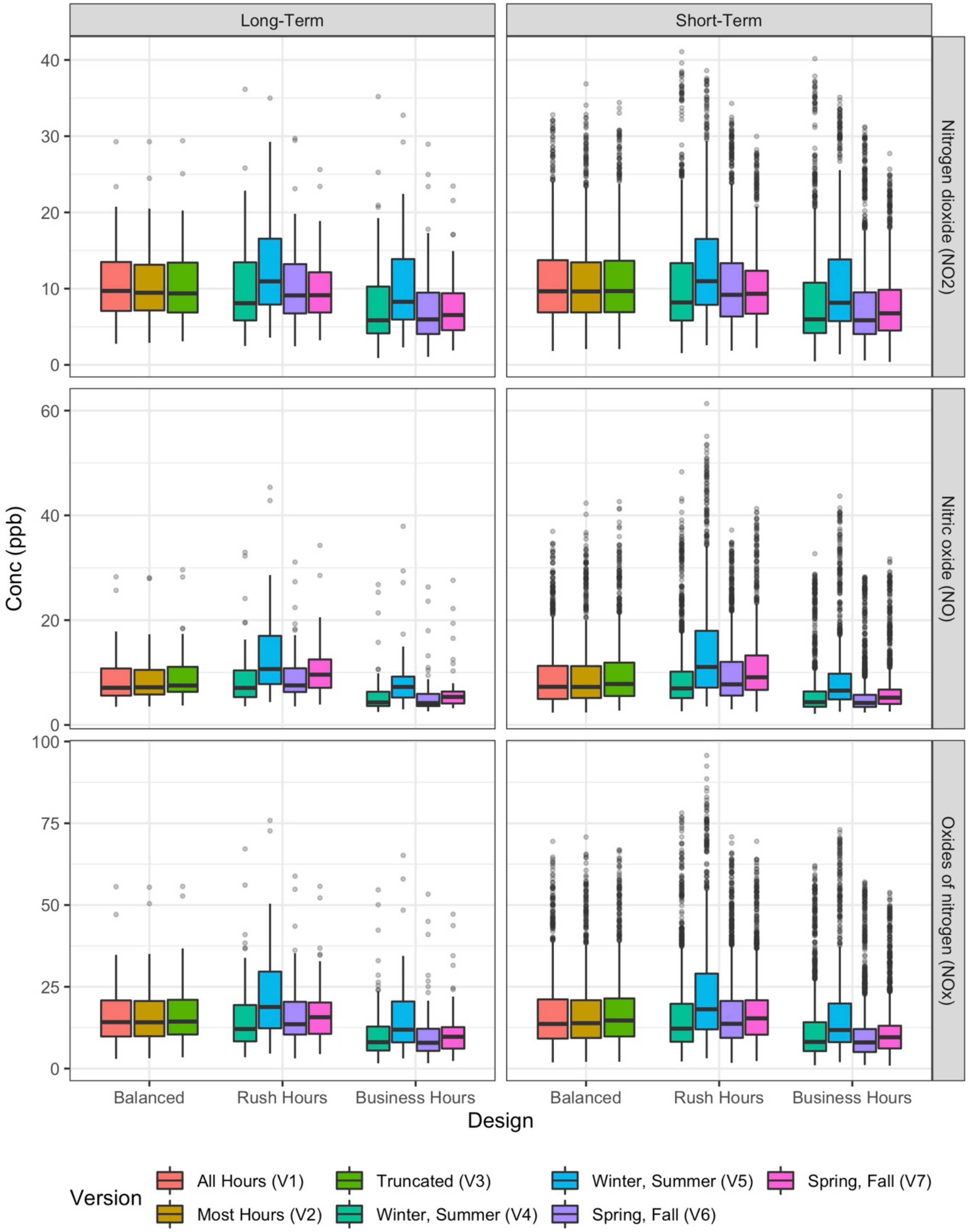

*Figure S7. Annual average site concentration estimates for different pollutants and design versions. N=30 campaigns per design version x 69 sites for short-term approaches; N = 1 campaign per design version x 69 sites for long-term approaches. Short-term approaches appear to be more variable (less precise), in large part because all 30 campaigns are represented in the boxplots.*

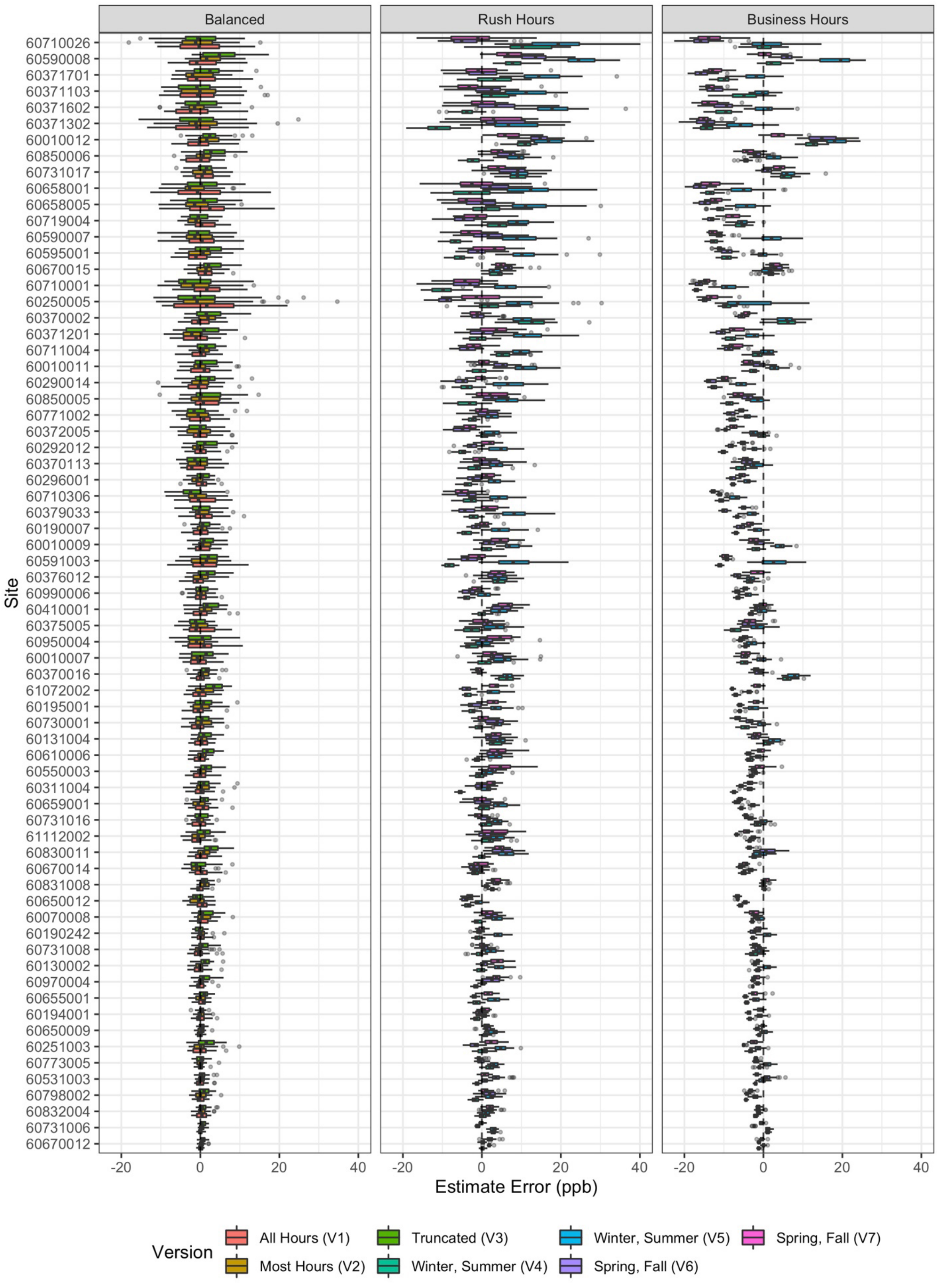
Balanced
Rush Hours
Business Hours
Site
60710026
60590008
60371701
60371103
60371602
60371302
60010012
60850006
60731017
60658001
60658005
60719004
60590007
60595001
60670015
60710001
60250005
60370002
60371201
60711004
60010011
60290014
60850005
60771002
60372005
60292012
60370113
60296001
60710306
60379033
60190007
60010009
60591003
60376012
60990006
60410001
60375005
60950004
60010007
60370016
61072002
60195001
60730001
60131004
60610006
60550003
60311004
60659001
60731016
61112002
60830011
60670014
60831008
60650012
60070008
60190242
60731008
60130002
60970004
60655001
60194001
60650009
60251003
60773005
60531003
60798002
60832004
60731006
60670012
-20
0
20
40
Estimate Error (ppb)
Version
All Hours (V1)
Most Hours (V2)
Truncated (V3)
Winter, Summer (V4)
Winter, Summer (V5)
Spring, Fall (V6)
Spring, Fall (V7)

*Figure S8. Site-specific NOx estimate error for short-term designs (N = 30 campaigns) as compared to the true estimates (long-term Balanced Design Version 1). All sites are included. Sites are arranged by the true NOx average, with higher concentrations higher up.*

#### 4 Model Predictions

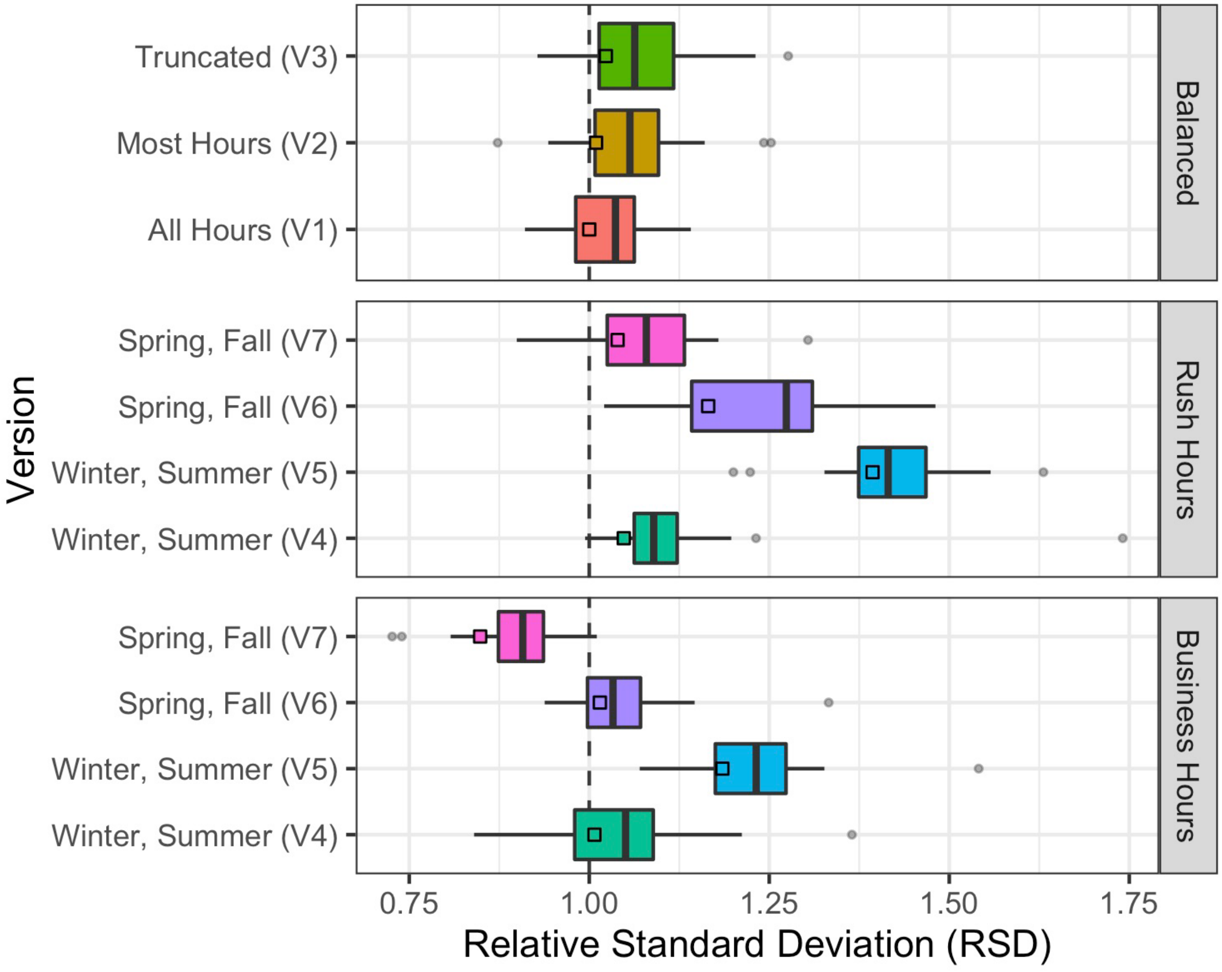

*Figure S9. Variation of predictions across 69 sites by design relative to the gold standard predictions (relative standard deviation [RSD]). Boxplots are for short-term approaches (30 campaigns), squares are for long-term approaches (1 campaign). Values of 1 indicate that design predictions have the same standard deviation as the gold standard model predictions.*

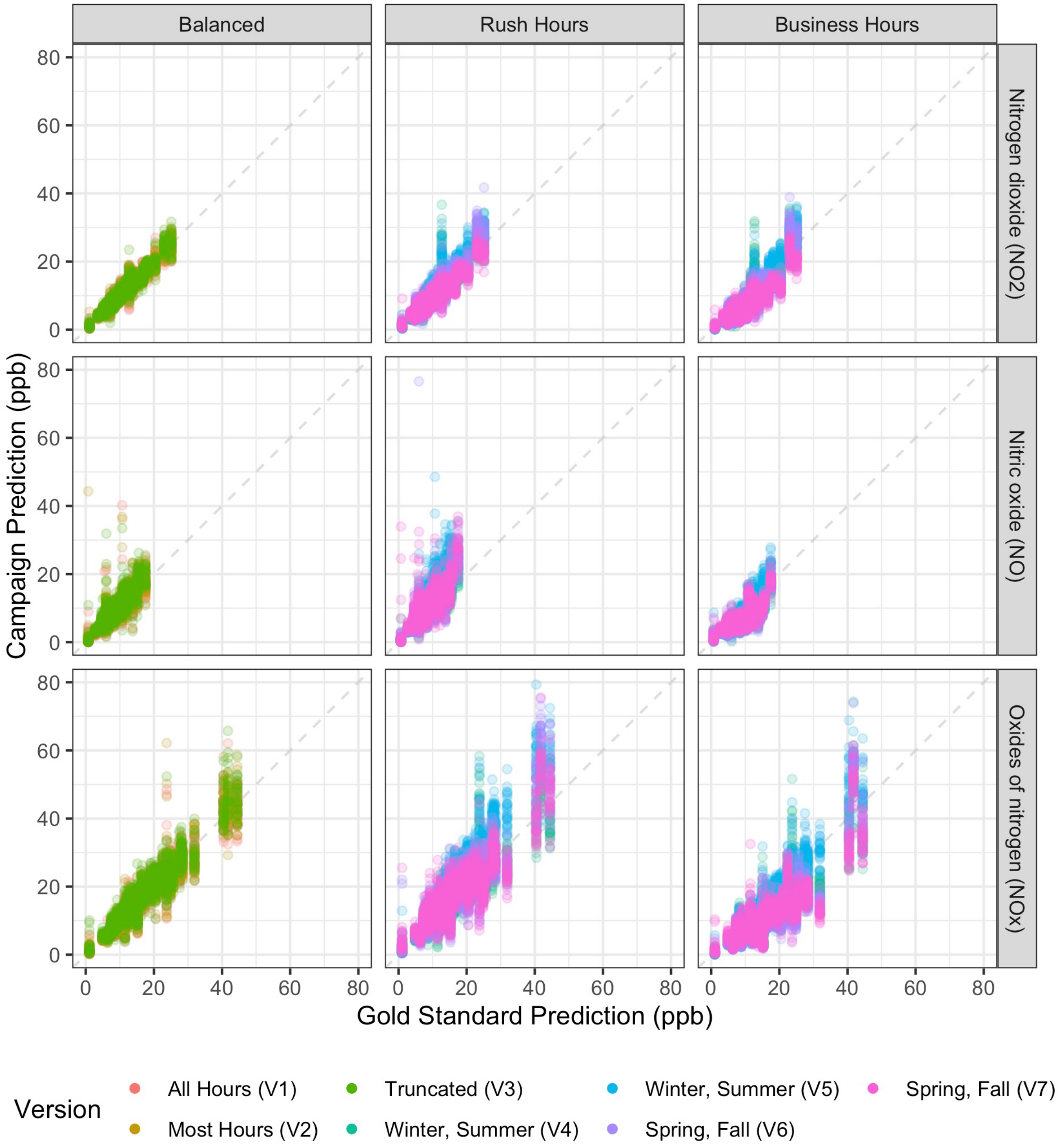

*Figure S10. Scatterplot of cross-validated short-term predictions for 30 campaigns vs the gold standard predictions for NOx, NO, and NO2. Showing predictions below 80 ppb for clarity (see SI Figure S12 and Table S6 for predictions excluded).*

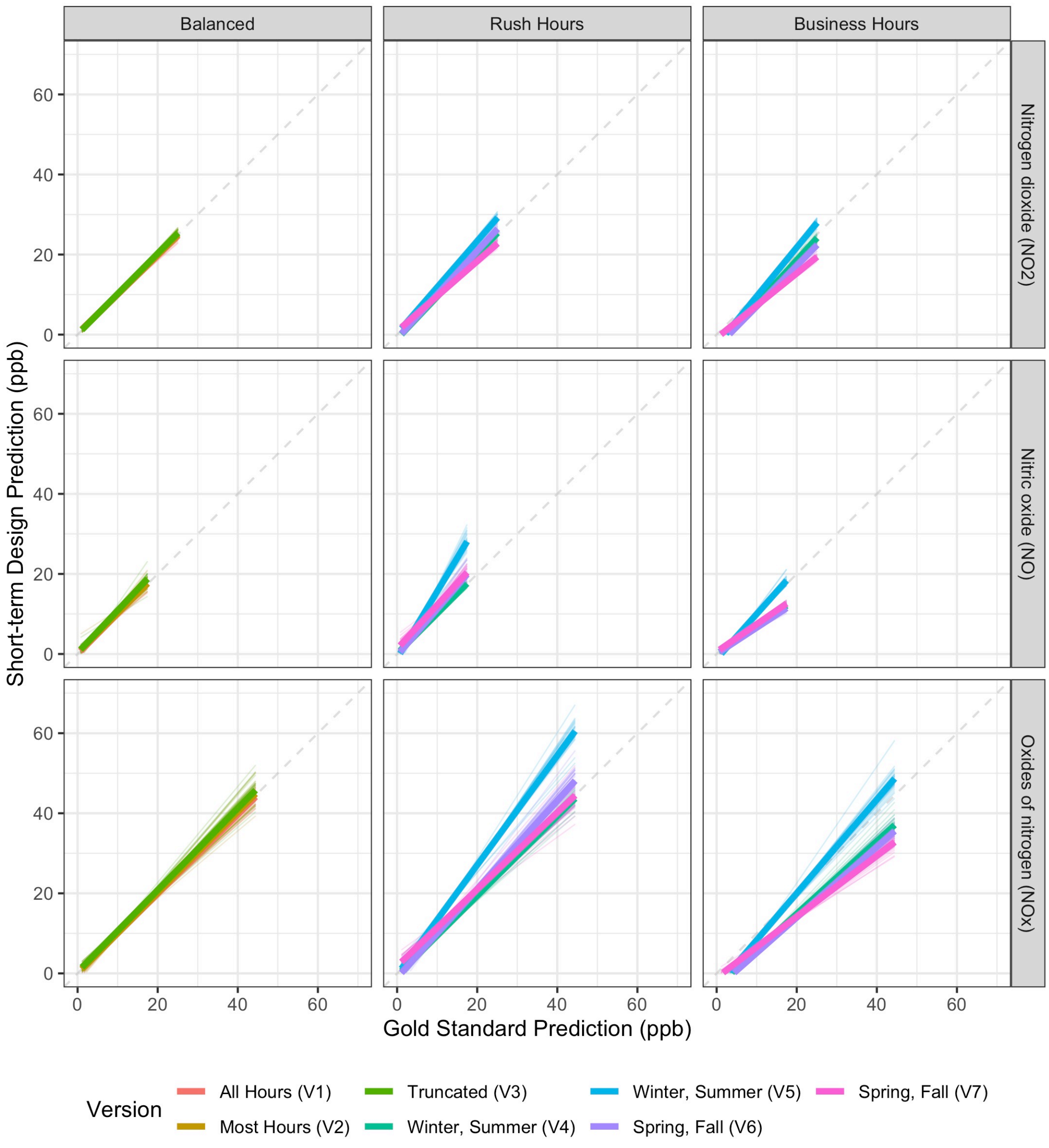

*Figure S11. Best fit lines of cross-validated short-term predictions for 30 campaigns vs the gold standard predictions for NOx, NO, and NO2.*

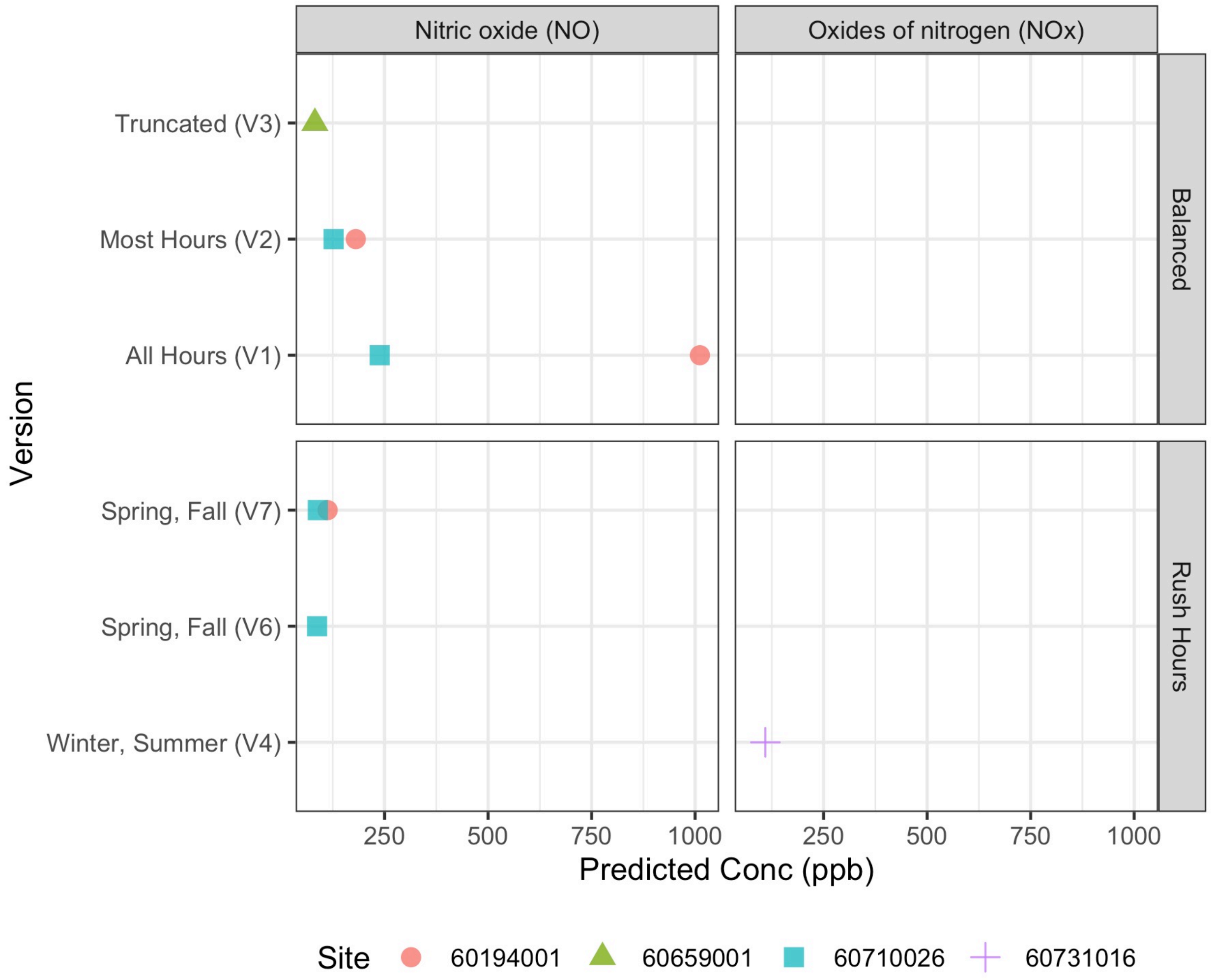

*Figure S12. Predictions above 80 ppb excluded from prediction plots, if noted.*

*Table S6. Predictions above 80 ppb excluded from prediction plots, if noted*[1]

| Parameter Name | Design | Version | N | Prediction (ppb) |
|---|---|---|---|---|
| Oxides of nitrogen (NOx) | Rush Hours | Winter, Summer (V4) | 1 | 109 |
| Nitric oxide (NO) | Balanced | All Hours (V1) | 2 | 238, 1012 |
| Nitric oxide (NO) | Balanced | Most Hours (V2) | 2 | 127, 181 |
| Nitric oxide (NO) | Balanced | Truncated (V3) | 1 | 82 |
| Nitric oxide (NO) | Rush Hours | Spring, Fall (V6) | 1 | 87 |
| Nitric oxide (NO) | Rush Hours | Spring, Fall (V7) | 2 | 89, 113 |

[1] N is the number of predictions.

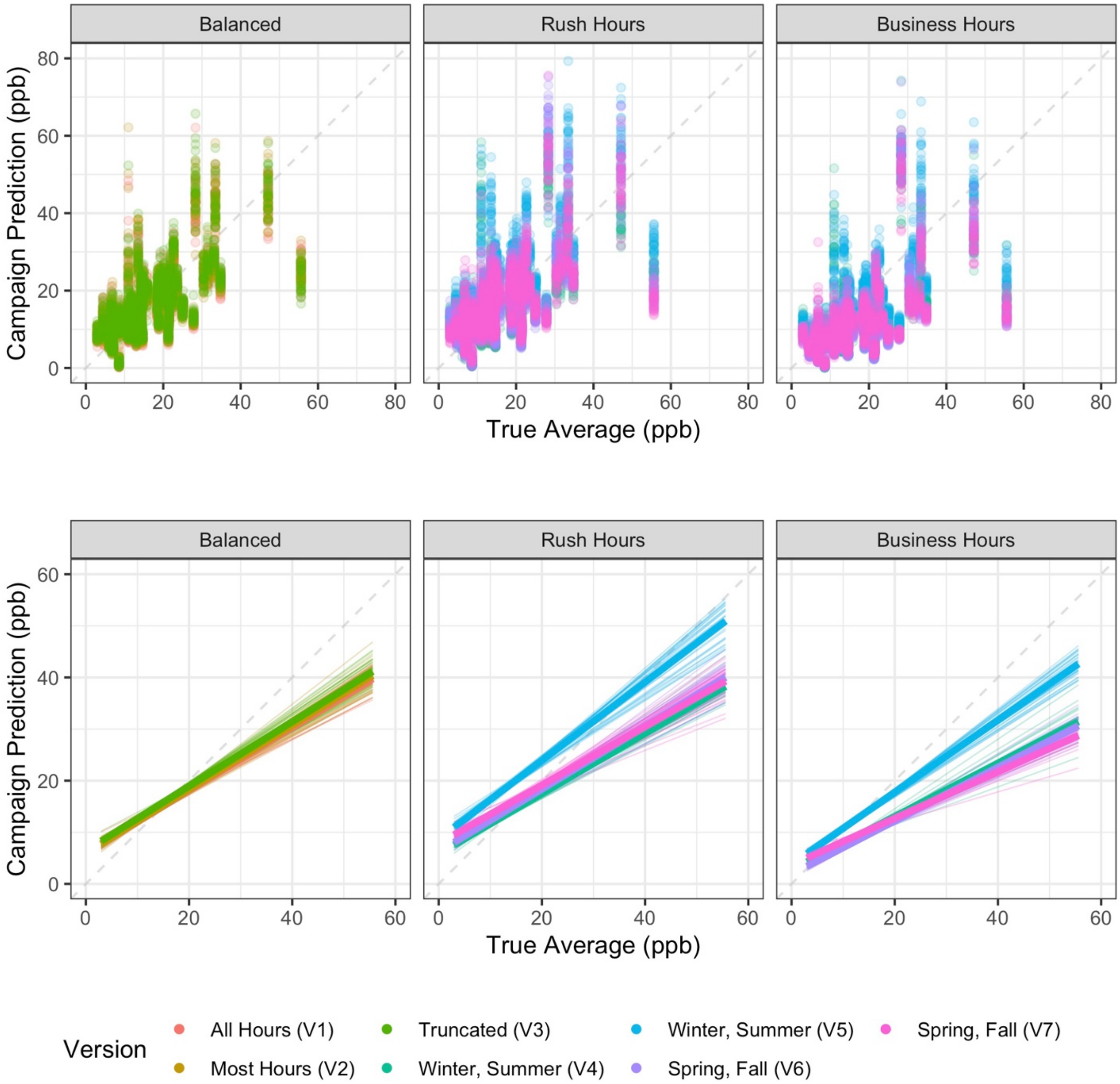

*Figure S13. Scatterplots and best fit lines of cross-validated short-term predictions for 30 campaigns vs true average concentrations for NOx. Thin transparent lines are individual campaigns, colored by design version; thicker lines are the overall version trend. (One prediction is excluded for clarity from the Rush Hours Version 4 scatterplot at x=24 ppb, y=109 ppb [site 60731016] but is included in the line plots).*

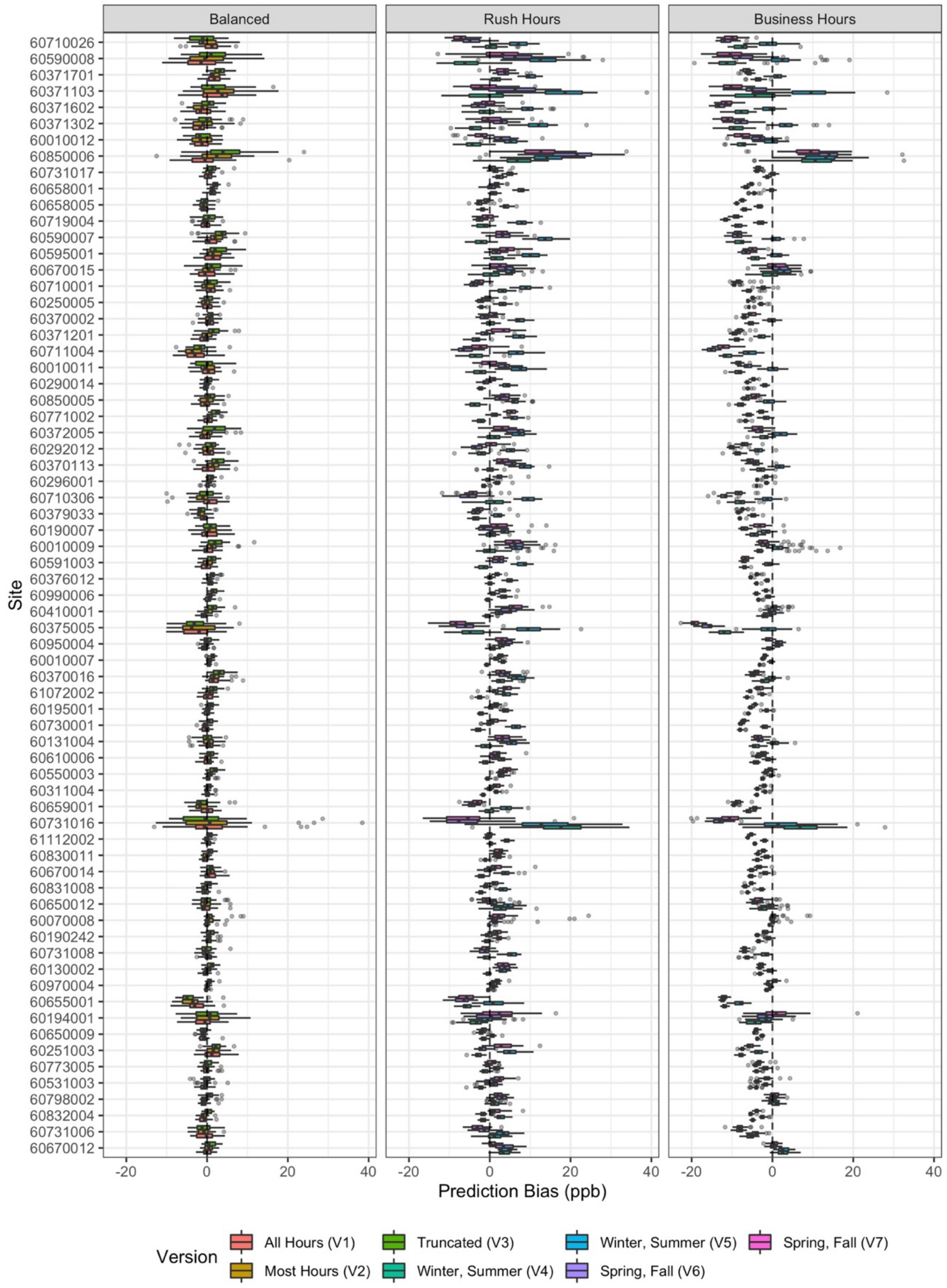

*Figure S14. Site-specific NOx prediction biases for short-term designs (N = 30 campaigns) as compared to the gold standard (long-term Balanced Design Version 1) predictions for all sites. Sites are arranged by the true NOx measurement, with higher concentration sites higher up. One prediction bias for site 60731016 is excluded (86 ppb for Rush Hours Version 4) for clarity.*

*Table S7. Distribution of prediction bias for short-term approaches relative to the gold standard predictions[1]*

| Parameter Name | Design | N | Min | Q01 | Median | IQR | Q99 | Max |
|---|---|---|---|---|---|---|---|---|
| Oxides of nitrogen (NOx) | Balanced | 6,210 | -13.1 | -6.9 | 0.2 | 2.4 | 7.9 | 38 |
| Oxides of nitrogen (NOx) | Rush Hours | 8,280 | -16.6 | -8.9 | 1.2 | 5.2 | 18.4 | 86 |
| Oxides of nitrogen (NOx) | Business Hours | 8,280 | -22.7 | -15.2 | -3.8 | 5.3 | 12.8 | 33 |
| Nitric oxide (NO) | Balanced | 4,590 | -10.5 | -3.8 | 0.1 | 1.7 | 7.2 | 1,006[2] |
| Nitric oxide (NO) | Rush Hours | 6,120 | -7.9 | -3.8 | 1.3 | 3.4 | 13.1 | 107 |
| Nitric oxide (NO) | Business Hours | 6,120 | -10.6 | -7 | -1.8 | 3 | 4.2 | 10 |
| Nitrogen dioxide (NO2) | Balanced | 6,300 | -6.9 | -3 | 0.1 | 1.1 | 3.5 | 11 |
| Nitrogen dioxide (NO2) | Rush Hours | 8,400 | -8.2 | -4.7 | 0.1 | 2.3 | 6.6 | 24 |
| Nitrogen dioxide (NO2) | Business Hours | 8,400 | -11.5 | -7.5 | -2.2 | 2.7 | 6.4 | 19 |

[1] N = the number of sites x 30 campaign repetitions x the number of versions per design
[2] This maximum is the result of a very large outlier prediction

### 5 Model Assessment

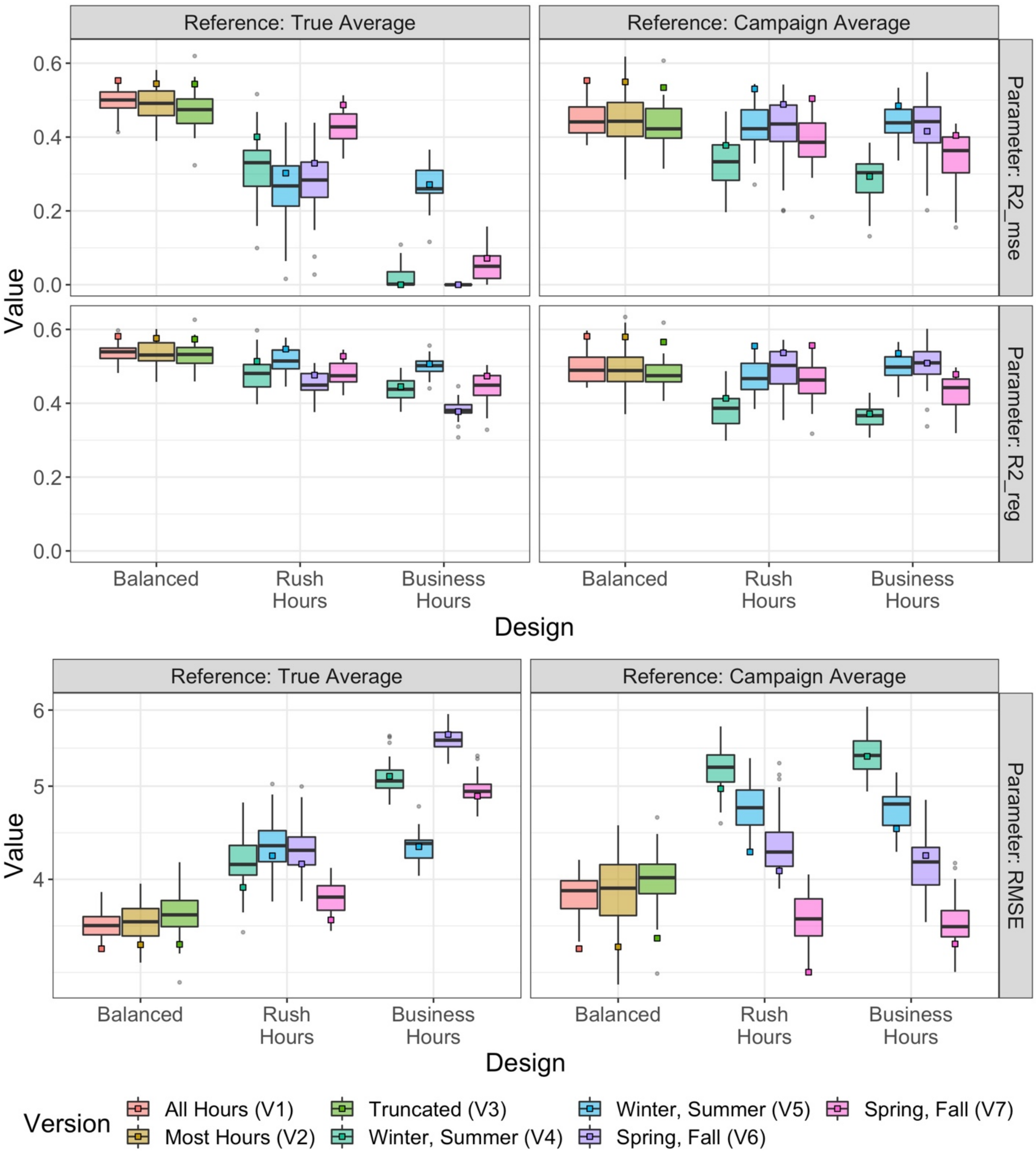

*Figure S15. $NO_2$ Model performances ($R^2_{MSE}$, $R^2_{reg}$, and RMSE), as determined by each campaign's cross-validated predictions relative to: a) the true averages (long-term Balanced Version 1), and b) its campaign averages. Boxplots are for short-term approaches (30 campaigns),while squares are for long-term approaches (1 campaign).*

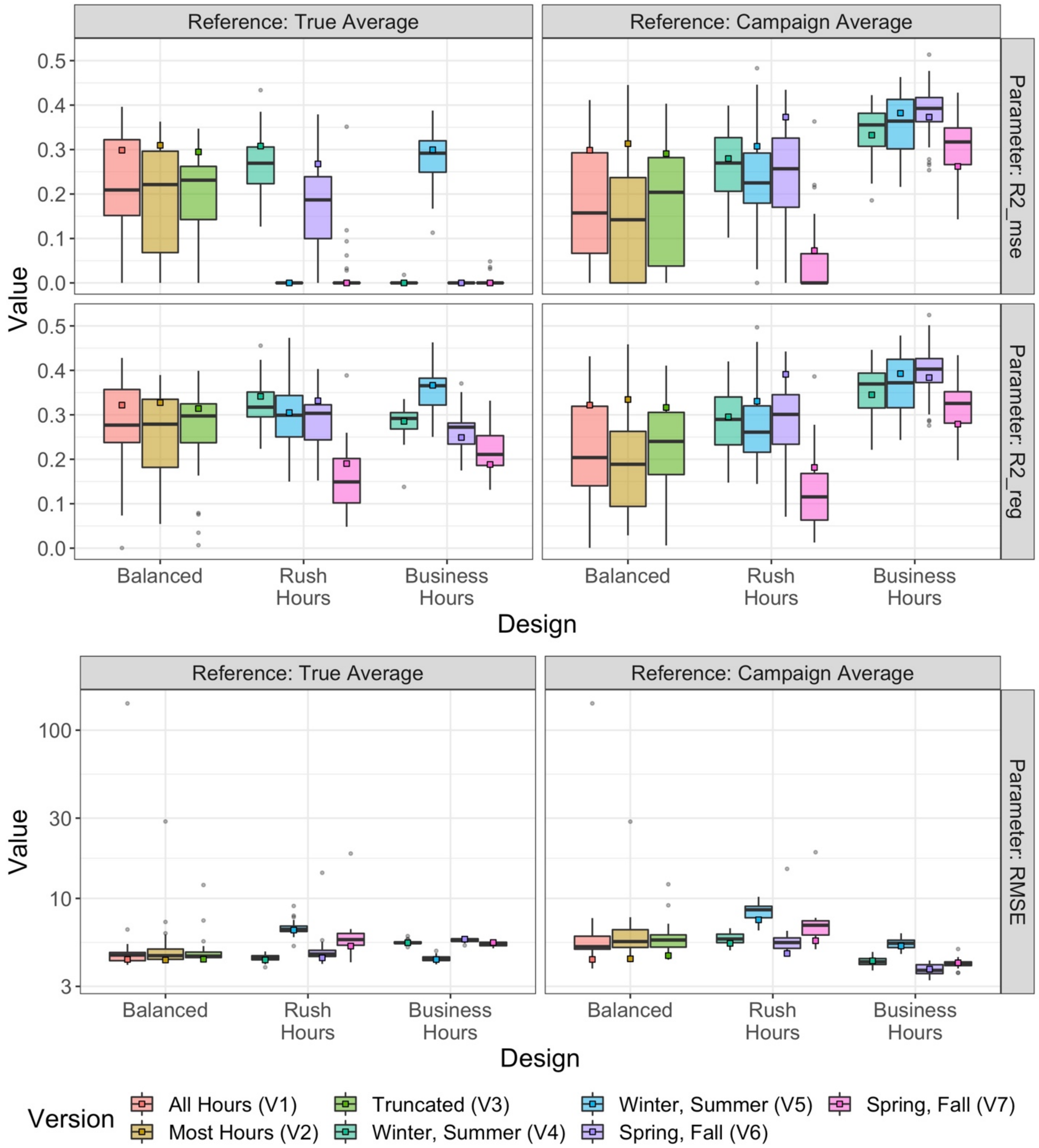

*Figure S16. NO Model performances ($R^2_{MSE}$, $R^2_{reg}$, and RMSE), as determined by each campaign's cross-validated predictions relative to: a) the true averages (long-term Balanced Version 1), and b) its respective campaign averages. Boxplots are for short-term approaches (30 campaigns),while squares are for long-term approaches (1 campaign). A few influential outliers influenced these performance statistics more so than for NOx and $NO_2$.*

### 6 Sensitivity Analyses

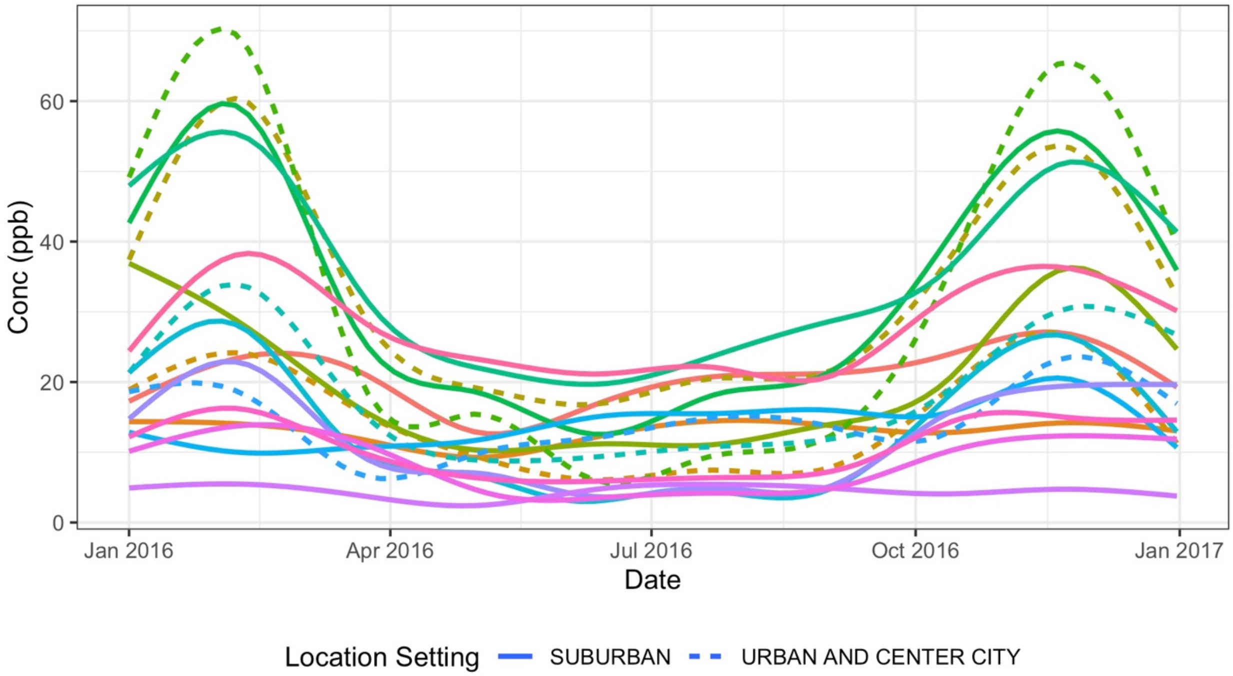

*Figure S17. Concentration trends for NOx at AQS sites included in the Los Angeles-San Diego analysis (N=17). Colored smooth lines are individual sites.*

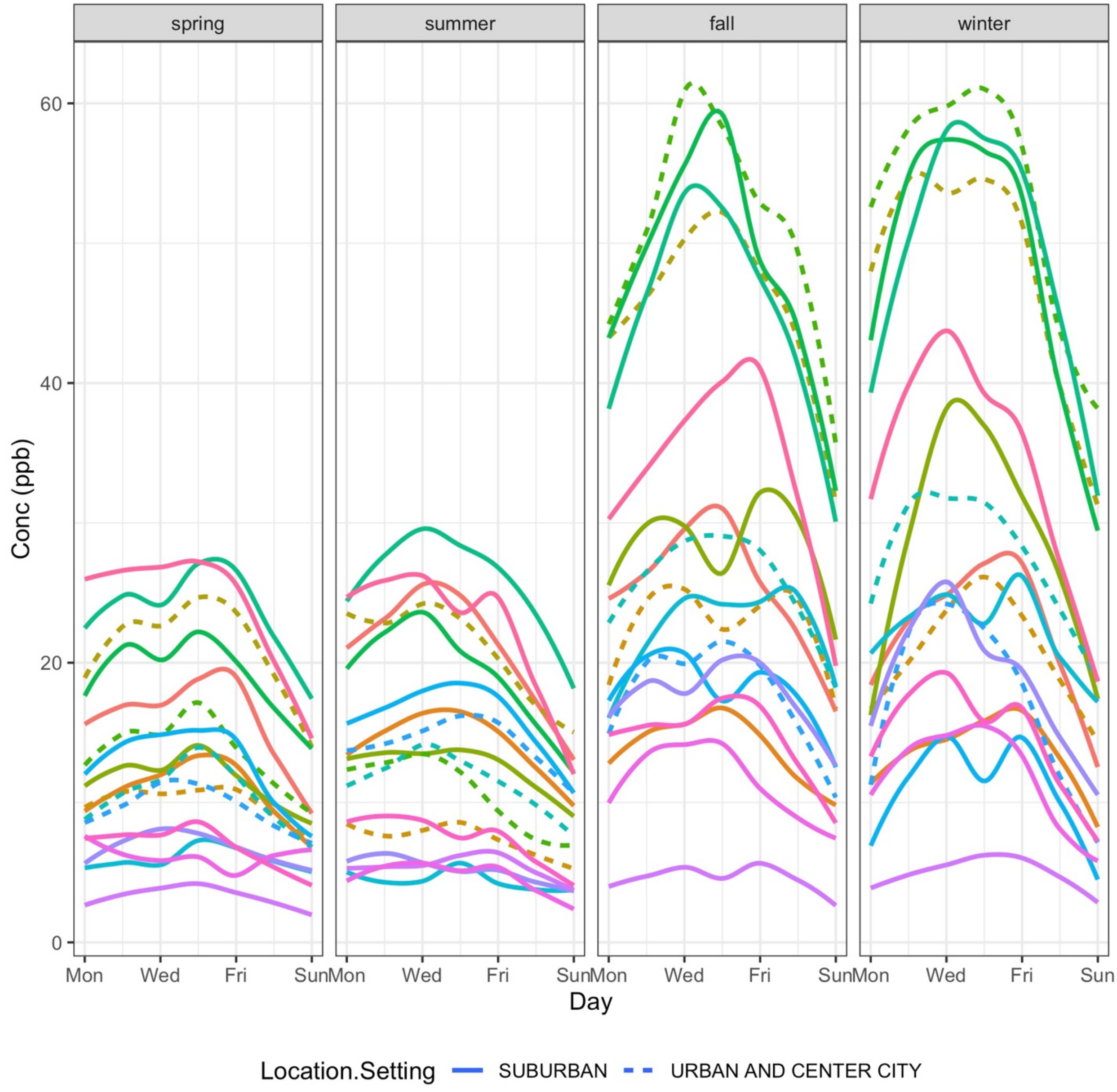

*Figure S18. Concentration trends for NOx at AQS sites included in the Los Angeles-San Diego analysis (N=17) by day and season. Colored smooth lines are individual sites.*

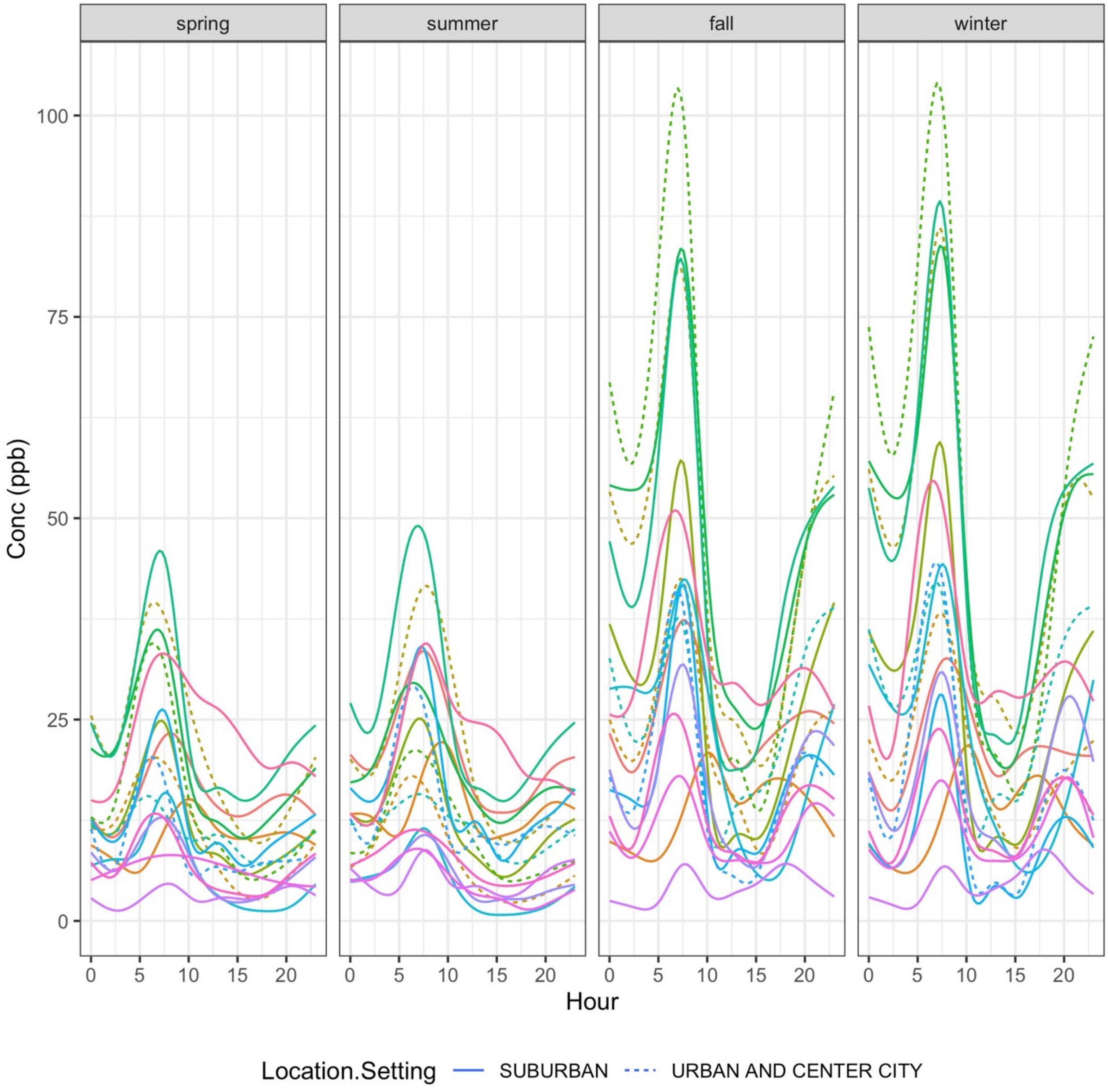

*Figure S19. Concentration trends for NOx at AQS sites included in the Los Angeles-San Diego analysis (N=17) by hour and season. Colored smooth lines are individual sites.*
